# Global impact and cost-effectiveness of one-dose versus two-dose human papillomavirus vaccination schedules: a comparative modelling analysis

**DOI:** 10.1101/2021.02.08.21251186

**Authors:** Kiesha Prem, Yoon Hong Choi, Élodie Bénard, Emily A Burger, Liza Hadley, Jean-François Laprise, Mary Caroline Regan, Mélanie Drolet, Stephen Sy, Kaja Abbas, Allison Portnoy, Jane J Kim, Marc Brisson, Mark Jit

## Abstract

**Background:** To eliminate cervical cancer as a public health problem, the World Health Organization currently recommends routine vaccination of adolescent girls with two doses of the human papillomavirus (HPV) vaccine before sexual initiation. However, many countries have yet to implement HPV vaccination because of financial or logistical barriers to delivering two doses outside the infant immunisation programme.

**Methods:** Using three independent HPV transmission models, we estimated the long-term health benefits and cost-effectiveness of one-dose versus two-dose HPV vaccination, in 188 countries, assuming that one dose of the vaccine gives either a shorter duration of full protection (20 or 30 years) or lifelong protection but lower vaccine efficacy (e.g., 80%) compared to two doses. We simulated routine vaccination with the 9-valent HPV vaccine in 10-year-old girls at 80% coverage for the years 2021–2120, with a one-year catch-up campaign of 11–14-year-old girls at 80% coverage in the first year of the programme.

**Results:** Over the years 2021–2120, one-dose vaccination at 80% coverage was projected to avert 112.9 million (range of medians: 75.8–176.2) and 148.0 million (111.6–187.6) cervical cancer cases assuming one dose of the vaccine confers 20 and 30 years of protection, respectively. Should one dose of the vaccine provide lifelong protection at 80% vaccine efficacy, 155.2 million (143.7–170.3) cervical cancer cases could be prevented. Around 65 to 889 additional girls would need to be vaccinated with the second dose to prevent one cervical cancer case, depending on the epidemiological profiles of the country. Across all income groups, the threshold cost for the second dose was low: from 0.85 (0.07–3.82) USD in low-income countries to 18.08 (−3.62–85.64) USD in high-income countries, assuming one-dose confers 30-year protection.

**Conclusions:** Results were consistent across the three independent models and suggest that one-dose vaccination has similar health benefits to a two-dose programme while simplifying vaccine delivery, reducing costs, and alleviating vaccine supply constraints. The second dose may be cost-effective if there is a shorter duration of protection from one dose, cheaper vaccine and vaccination delivery strategies, and high burden of cervical cancer.

## Background

Cervical cancer is the fourth leading cause of cancer mortality among women globally with an estimated 570 000 new cases and 311 000 deaths in 2018, with the majority of deaths occurring in low- and middle-income countries (LMICs) (1). Persistent infection with high-risk genotypes of human papillomavirus (HPV) is a necessary precursor of cervical cancer.

Primary prevention of cervical cancer is available with four highly efficacious prophylactic vaccines—two 2-valent, one 4-valent, one 9-valent—that are currently licensed for protection against HPV infection (2–5). All protect against the two most carcinogenic HPV types, 16 and 18, which are responsible for 70% of cervical cancer cases globally (6–8). Some additionally protect against HPV types 6 and 11, which do not cause cancer but are responsible for most cases of anogenital warts, and against other high-risk types such as HPV 31, 33, 45, 52, and 58 (either directly or through cross-protection), which have been linked to a further 20% of cervical cancer cases (6–8).

Multiple analyses including the global Papillomavirus Rapid Interface for Modelling and Economics (PRIME) model developed in collaboration with the World Health Organization (WHO) (9,10) have found HPV vaccination to be cost-effective in almost all countries. The HPV vaccines were initially administered as a three-dose regimen over six months. In 2014, the WHO Strategic Advisory Group of Experts on Immunization reviewed the evidence for dose reduction and recommended a two-dose regimen for individuals below 15 years of age (11). With the availability of vaccines and screening tests that allow detection of both high-risk HPV types and neoplasias that are precursors to cervical cancer, the Secretary-General of WHO has called for global elimination of cervical cancer as a public health problem, i.e., achieving the measurable global targets set by WHO (12). Current WHO guidelines recommend that all countries vaccinate females aged 9–14 years against HPV (13).

Although some of these vaccines have been licensed for more than a decade, LMICs with the highest incidence of cervical cancer are disproportionately less likely to introduce the HPV vaccine into their routine immunisation programmes (9,14–16). High vaccine procurement and delivery costs coupled with logistical constraints surrounding the delivery of a two-dose regimen outside the infant vaccination schedule has hampered vaccine introduction and uptake (17). Despite the financial support of Gavi, the Vaccine Alliance, many LMICs have yet to introduce HPV vaccines into their routine programmes (18,19). Since 2017, constrained supply of the 4-valent and 9-valent HPV vaccines has further delayed vaccine introductions in many countries (20,21). Moreover, physical distancing measures such as school closures and national lockdowns in response to the current COVID-19 pandemic (22) have caused eligible populations to miss doses of HPV vaccine (21).

These financial, logistical, and supply constraints have motivated research into one-dose vaccination schedules. If proven effective, one-dose HPV vaccination would simplify vaccine delivery and lower costs of national vaccination programmes (18,23). It could also expedite the introduction of HPV vaccines into national immunisation schedules for LMICs, potentially protecting many more females against cervical cancer (19).

Evidence is emerging from immunogenicity trials, post-hoc analyses of efficacy trials, and post-licensure observational studies to suggest that one dose of the HPV vaccine may provide a high level of protection against incident and persistent HPV infections. A systematic review of participants in six clinical trials who received only one dose of HPV vaccination, because they did not complete their allocated schedules, suggests that this schedule may be as effective as two doses in preventing HPV infection in up to seven years of follow-up (24). However, evidence on the non-inferior efficacy of a single-dose schedule from participants randomised to receive one dose has yet to emerge (expected in 2025). Furthermore, antibody titres in immunogenicity trials were lower than in those receiving two or three doses. While inferior antibody titres may not necessarily translate to inferior protection, at this point, there is still uncertainty about the efficacy and durability of one-dose vaccination.

Additionally, in the event that one-dose vaccination protection is slightly worse than two or three doses, populations may still be almost as well protected through indirect (herd) protection. Such effects can be examined using HPV transmission dynamic models. To date, model-based analyses set in the United Kingdom (UK) (25), the United States (US), and Uganda (26,27) suggest that one-dose schedules would be cost-effective and would prevent almost as many cancers as two-dose or three-dose schedules if one dose confers at least 20 years of protection or has at least 80% efficacy against HPV 16/18 infection.

In this paper, we compare the impact and cost-effectiveness of one-dose versus two-dose vaccination in 188 countries, assuming that one dose of the vaccine gives either shorter duration of protection or lower vaccine efficacy compared to two doses. We use a hybrid approach: firstly, we consider the age-specific impact that HPV vaccines may have using the results of multiple independent HPV transmission dynamic models, and secondly, extrapolate these effects to the remaining countries in the world using data on population demographics and cervical cancer burden synthesised in a single model (PRIME).

## Methods

To assess the extent to which one-dose HPV vaccination schedules will provide similar protection and be cost-effective compared to two doses, we compared the impact of three different strategies: (1) no HPV vaccination; (2) a one-dose HPV vaccination schedule in which we assume that one dose of the HPV vaccine confers either 20 or 30 years of full protection or 80% vaccine efficacy (VE) over the lifetime; and (3) a two-dose HPV vaccination schedule in which two doses of the vaccine would provide lifetime protection at 100% VE. The minimum duration of protection in the waning scenarios for one-dose reflects the availability of over 10 years of data from various studies—ESCUDDO trial (28,29), IARC India post-randomisation analysis (30)—that do not show any evidence of waning of either clinical or immunological protection (31).

Fig 1 provides an overview of the data sources and key steps of the modelling framework described in the following sections. We synthesised the long-term population-wide impact of HPV vaccination on cervical cancer incidence by age and time predicted by three published transmission dynamic models: (i) the Public Health England (PHE) model, a compartmental dynamic model set in the UK (32); (ii) the HPV-ADVISE model, an individual-based dynamic model set in Uganda, Nigeria, India, Vietnam (27,33), and Canada (34,35); and (iii) the Harvard model, a hybrid model that links two individual-based models, set in the US, Uganda, El Salvador, and Nicaragua (14,36). In total, we combined results from 10 model-country scenarios. The models have been extensively reviewed and used to inform vaccine policy (including by the UK’s Joint Committee on Vaccination and Immunisation (37), the World Health Organization’s Strategic Advisory Group of Experts on Immunization (11,38,39) and the US Advisory Committee on Immunization Practice (40–43). The models stratify population by age, gender, and sexual activity-based risk group, as well as screening behaviour-based risk group in the HPV-ADVISE and Harvard models. They capture HPV natural history and disease, as well as HPV transmission as informed by country-specific sexual behaviour surveys. More details about the models can be found in the Supplementary Materials. For the scenarios where one dose confers a shorter duration of protection (i.e., 20 or 30 years), we assume 100% VE, as suggested by clinical trial populations (24,28,30,31). We modelled routine annual vaccination with the 9-valent vaccine in 10-year-old girls to begin in 2021 and run uninterrupted until 2120. We also included catch-up vaccination of girls aged 11–14 years in the first year of the programme. Throughout, vaccine coverage was assumed to be 80%. In sensitivity analyses, we investigated the impact of a one-dose vaccination schedule with a bivalent vaccine (Supplementary Materials).

**Figure 1.**
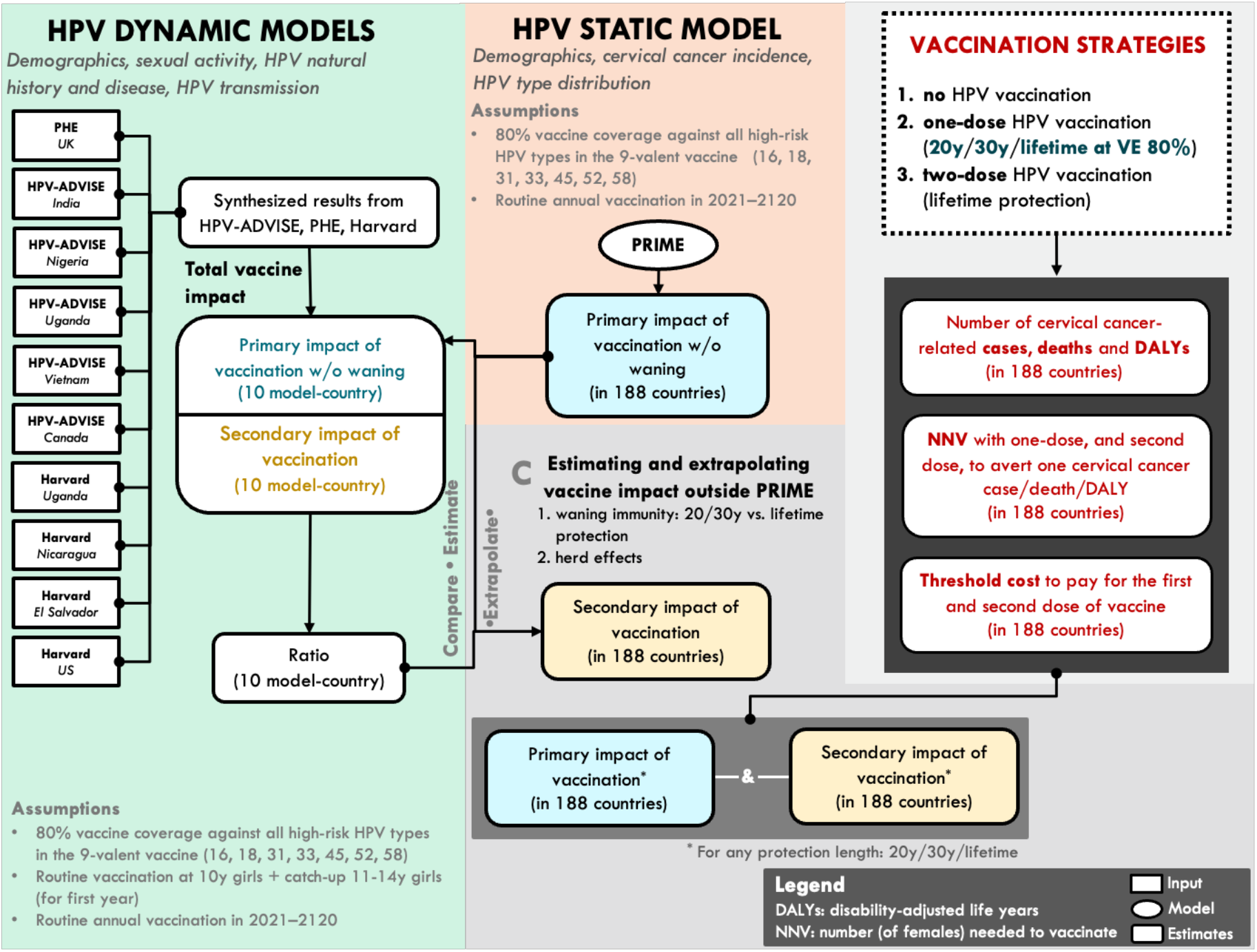
Overview of the data sources and the key steps of the modelling. To compare the impact and cost-effectiveness of one-dose versus two-dose vaccination in 188 countries, we adopted a hybrid approach. First, we synthesised the age-specific impact of HPV vaccines of three published transmission dynamic models—PHE, HPV-ADVISE, Harvard—from 11 model-country settings. Second, we derived the primary impact of vaccination using a static model (PRIME). Third, we extrapolated the primary and secondary effects to the remaining countries in the world. Fourth, we measured and compared population-level impact (e.g., cervical cancers averted, number of females needed to be vaccinated, threshold costs of the first and second dose of the vaccine) for three vaccine strategies: no HPV vaccination (the counterfactual); a one-dose HPV vaccination schedule in which we assume that one dose of the vaccine provides either a shorter duration of protection (20 or 30 years) or lower vaccine efficacy (i.e., 80%) compared to two doses; and a two-dose HPV vaccination schedule in which two doses of the vaccine provides lifetime protection.

Using PRIME, we then estimated the primary impact of a two-dose vaccination schedule, without herd effects and waning immunity, in 188 countries. Full details of PRIME, including model equations and updates, are available at (9,10). As PRIME is a static model, it cannot estimate herd effects, nor can it capture the effect of waning vaccine immunity. Here, we introduced a novel method which compares results from PRIME and the three dynamic models—PHE, HPV-ADVISE, Harvard—set in nine countries—UK, US, Canada, Nigeria, Uganda, India, Vietnam, El Salvador, and Nicaragua. We calculated the difference between cervical cancer incidence predicted by PRIME and each of the dynamic models to derive the secondary effects of vaccination, which is a combination of waning immunity (20/30-year duration vs lifetime protection and lower vaccine efficacy) and herd effects at every age and time-point. We then calculated the ratio of secondary to primary vaccine impact. By assuming that the primary impact of a vaccine (i.e., vaccine with lifetime protection and no herd effects) is different in every country as estimated by PRIME, we extrapolated the ratio (secondary to primary) to other countries to project the secondary effects of vaccination, using a similar approach as two comparative modelling analyses conducted by the WHO’s Cervical Cancer Elimination Modelling Consortium (14,44). A meta-analysis by Drolet and colleagues showed a significant decrease in the prevalence of HPV 16 and 18 among women aged 20–24 years (risk ratio [RR] 0.34, 95% CI 0.23–0.49) and 25–29 years (RR 0.63, 95% CI 0.41–0.97) (45). As most of the women in these age groups were unvaccinated, the meta-analysis found evidence of similar herd effects more than four years after the introduction of HPV vaccination.

Uncertainty in predictions was captured by generating multiple simulations from the three dynamic models representing different plausible parameter sets. For the PHE model, 100 runs were simulated from the best-fitting parameter sets to capture uncertainty in the duration of infection, duration of natural immunity, screening accuracy, the progression of cervical cancer, age-specific prevalence, and the number of sexual partners. For HPV-ADVISE, 1000 runs were simulated from 50 parameter sets that simultaneously fit country-specific behavioural and epidemiological data. These 50 parameter sets illustrate the uncertainty in sexual behaviour, HPV transmission, the natural history of HPV-related diseases, and screening. For the Harvard model that reflect two sexual behaviour settings (low- and high-HPV prevalence), 50 best-fitting dynamic transmission model parameter sets, capturing variations in genotype- and sex-specific transmission probability, and genotype- and sex-specific natural immunity, were propagated through four cervical carcinogenesis models that have been previously calibrated (i.e., fit) to the US, Uganda, El Salvador, or Nicaragua (14,36).

### Effectiveness and cost-effectiveness measures

For each country, we estimated the number of cervical cancer cases, deaths, and disability-adjusted life years (DALYs)—caused by HPV 16, 18, 31, 33, 45, 52, and 58—occurring under each scenario by age and time since vaccination in females born in the years 2011–2110 (Section 1.8 Supplementary materials). We then compared the impact of a one-dose schedule (giving 20/30 years protection or lifelong protection but at 80% initial VE) with no vaccination, and a two-dose schedule (giving lifetime protection at 100% VE) with a one-dose schedule. We calculated the number of females needed to vaccinate with one dose, and the number of females needed to give an additional (i.e., second) dose, to avert one cervical cancer case, death, or DALY. We also projected the threshold cost to pay for the first and second dose of vaccine, which is the maximum that could be paid for the first dose (compared to no vaccination) and second dose (compared to one dose only) for the incremental cost-effectiveness ratio to remain below country-specific gross domestic product (GDP) per capita (in 2017 USD). We used the GDP per capita estimates by the World Bank (46), but also considered a lower threshold, i.e., 30–40% and 60–65% of GDP per capita in low-income and middle- to high-income countries (47,48). The time horizon of the analysis was from 2021 to 2120; we accrued all health benefits of vaccination up to the end of the routine vaccination programme (i.e., the year 2120) or age 100 of all vaccinated cohorts, whichever came first. Using modelled results from the 10 model-country pairs, we projected the outcome measures in 188 countries and aggregated the results by World Bank income groups. After projecting the various measures of effectiveness and cost-effectiveness under the several vaccination scenarios, we compared the outcomes generated with results from the 10 model-country pairs. After projecting the various measures of effectiveness and cost-effectiveness under the vaccination scenarios, we compared the outcomes generated with results from the 10 model-country scenarios. We presented the results, aggregated by World Bank income groups (details in the Supplementary Materials), as the median (and 80% uncertainty intervals (UI)) from each of the model-country predictions. Both health outcomes and costs were discounted at 0% and 3% per year (49).

## Results

In 188 countries over the years 2021–2120, the models projected that routine annual vaccination of 10-year-old girls (plus a one-year catch-up campaign of girls aged 11–14 years) with one-dose of the 9-valent HPV vaccine at 80% coverage would avert 112.9 million (range of medians: 75.8–176.2) and 148.0 million (111.6–187.6) cervical cancer cases should one dose of the vaccine confer 20 and 30 years of protection, respectively (Fig 2; with the equivalent cumulative and discounted benefits figures in the Supplementary Materials). Under a scenario of one dose of the vaccine providing lifelong protection at 80% initial VE, the models predicted that 155.2 million (143.7–170.3) cervical cancer cases would be prevented (Fig 2). A one-dose schedule conferring 20 years of protection would avert 65.0% (range of medians: 45.8–91.8%) of the cases averted by the vaccination schedule providing lifelong protection at 100% VE (Fig 3). However, if the duration of protection increases to 30 years, a one-dose schedule would avert more cases at 87.9% (range of medians: 59.6–100%) of the cases averted by the vaccination schedule providing lifelong protection at 100% VE (Fig 3). Similarly, for the scenario where one dose of the vaccine provides lifelong protection but at lower VE (of 80%), most of the cases (88.3% (range of medians: 86.2–96.7%)) can still be averted (Fig 3).

**Figure 2.**
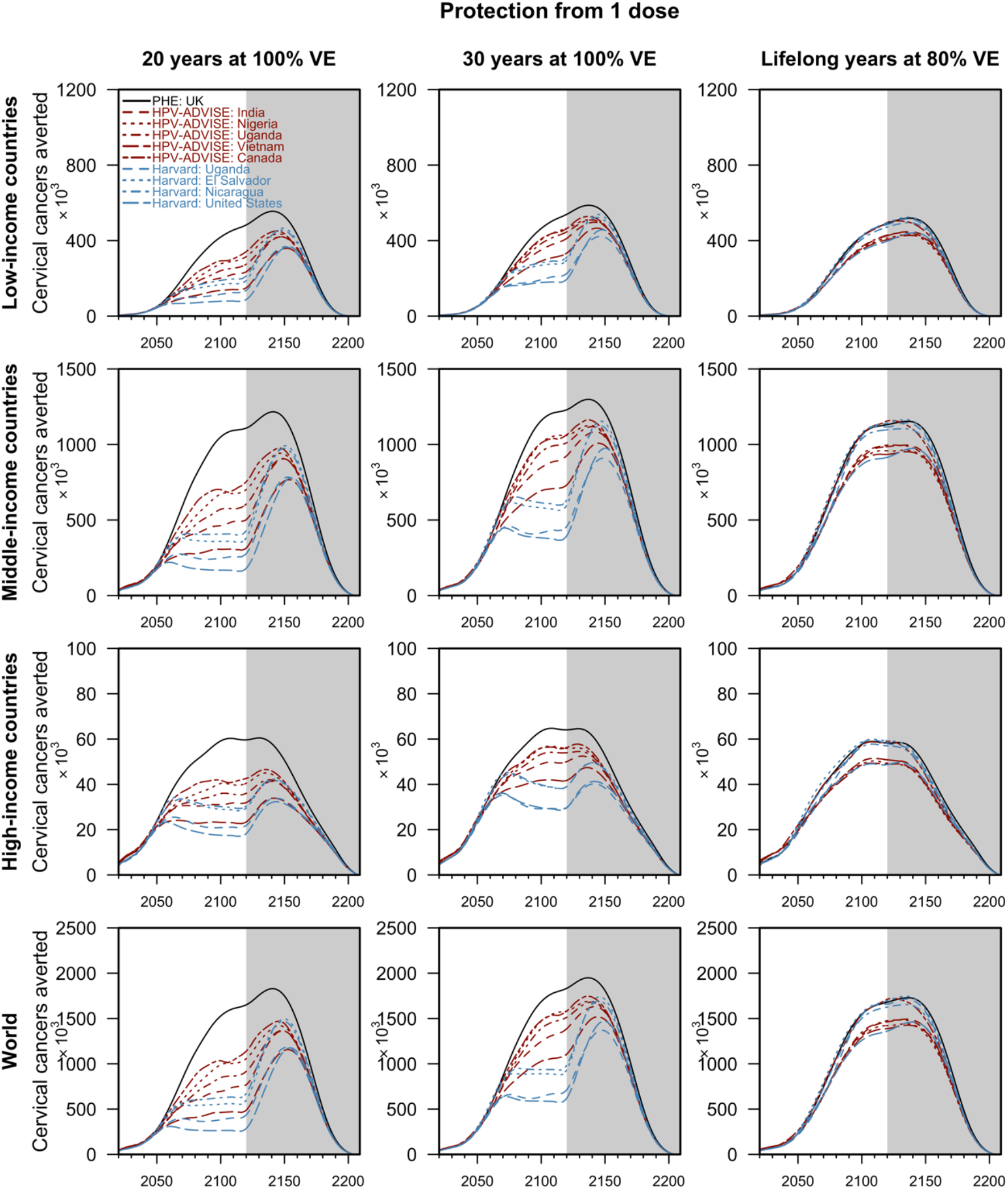
Cervical cancers averted by routine one-dose HPV vaccination by country income groups. The lines represent the median projections of the 10 model-country settings: the PHE model in black, HPV-ADVISE model-country pairs in red, and the Harvard model-country pairs in blue. The grey area corresponds to the additional cases averted in the vaccinated cohort after the 100 years of routine vaccination. Cancers averted (health outcomes) were discounted at 0%. Only cervical cancer caused by HPV 16, 18, 31, 33, 45, 52 and 58, which could be averted by the 9-valent HPV vaccine, were considered.

**Figure 3.**
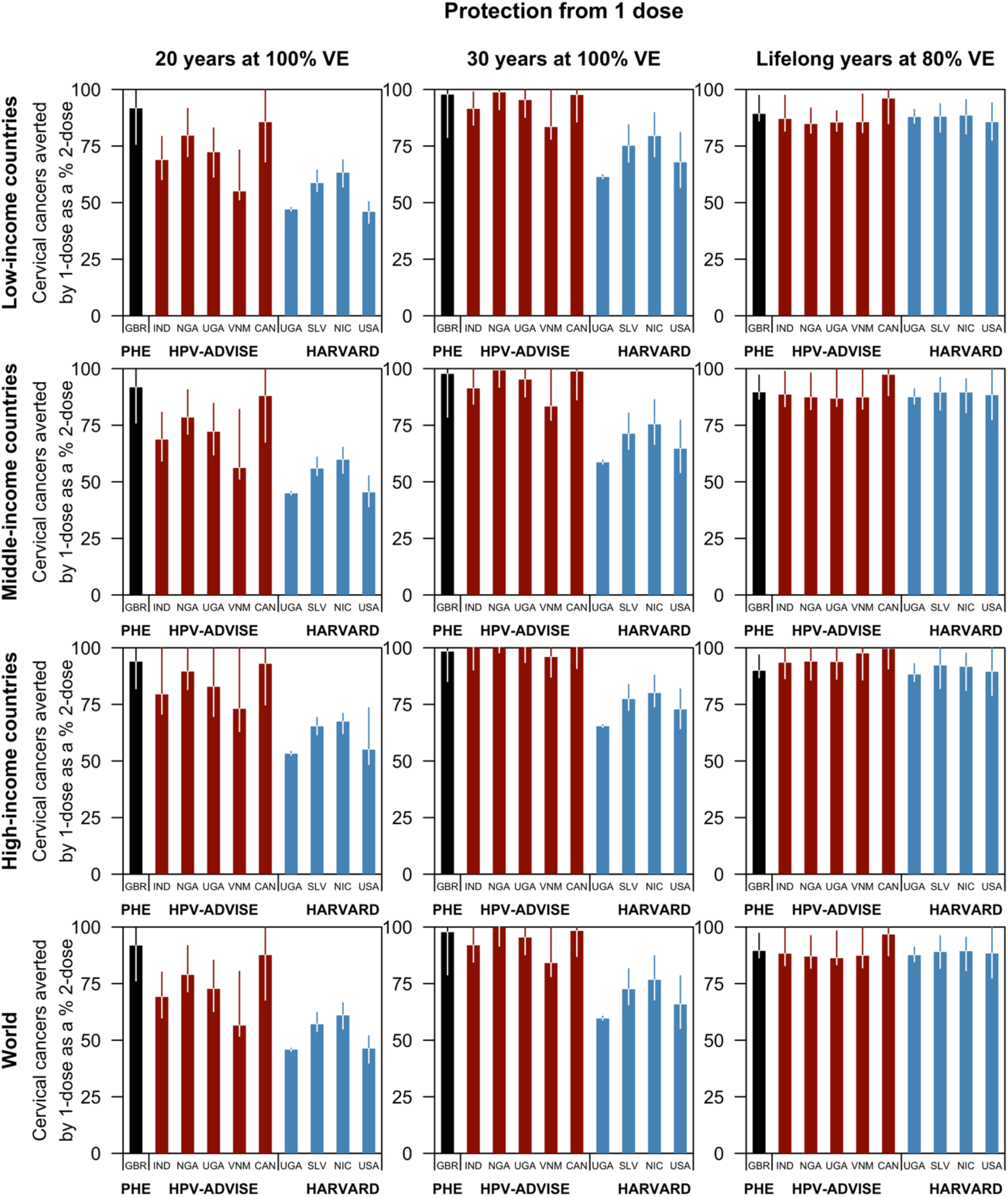
Cervical cancers averted by routine one-dose HPV vaccination as a proportion of cervical cancers averted by routine HPV vaccination programmes conferring lifelong protection at 100% vaccine efficacy. The median percentage (intervals: 10–90th percentile) of cancers averted by a one-dose schedule compared to a two-dose programme of the 10 model-country settings: the PHE model in black, HPV-ADVISE model-country pairs in red, and the Harvard model-country pairs in blue. Health outcomes were discounted at 0%. Only cervical cancer caused by HPV 16, 18, 31, 33, 45, 52 and 58, which could be averted by the 9-valent HPV vaccine, were considered.

Due to large disparities in age-standardised cervical cancer incidence across country income groups in 2021, the number of cases averted by routine vaccination programmes is higher in low-income countries (31.9 million (range of medians: 21.2–48.6), if one-dose confers 20 years of protection) than in high-income countries (4.8 million (range of medians: 3.5–6.9)). More cervical cancers could be averted if one dose of the vaccine confers a longer duration of protection, i.e., at 30 years or lifelong but lowered VE. Assuming waning of protection at 20 years (on average) after vaccination, the PHE model parameterised with data from the UK projected that a one-dose schedule could avert 91.8% (80%UI 76.2–99.8%) of the cases averted by a two-schedule vaccination schedule. However, the HPV-ADVISE and Harvard models, mostly parameterised with data from LMICs, projected that 61.0% (range of medians: 45.8–87.6%) could be averted (Fig 3).

The models consistently projected that fewer girls need to be vaccinated with the first dose to prevent one cervical cancer case in low-income countries (32 (range of medians: 23–51)) than middle-income (43 (range of medians: 31–69)) and high-income countries (83 (range of medians: 67–122)) if one-dose confers 20 years of protection (Fig 4A–C). However, variations across models were observed for the projections of the number of girls needed to be vaccinated with the second dose to prevent one cervical cancer case. Compared to the HPV-ADVISE and Harvard models, the PHE model projected that more girls need to be vaccinated with the second dose to avert one cervical cancer case when the protection from one dose of the vaccine wanes 20 years after vaccination (Fig 4A–D; 889 (80%UI 93–27 700) girls if one dose confers 20 years of protection). However, if one-dose confers lifelong protection but at lowered VE, the differences between the PHE, HPV-ADVISE and Harvard models decrease. When we discounted health outcomes, the model predicted that more girls need to be vaccinated to avert one cervical cancer case (Fig 4E–H).

**Figure 4.**
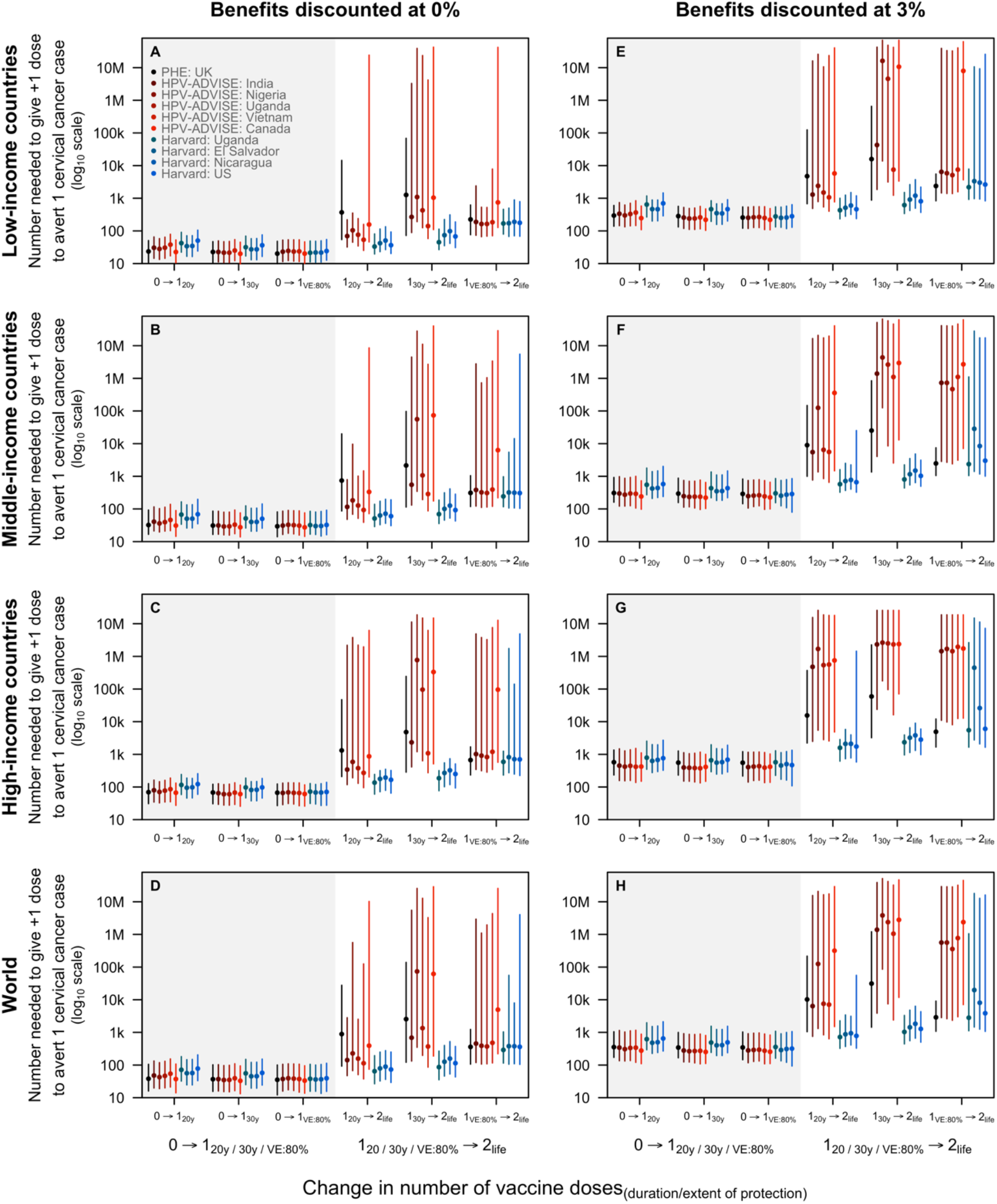
Number of girls needed to be vaccinated with the first and second dose to avert one additional cervical cancer case by income group. The lines represent the median projections of the 10 model-country settings: the PHE model in black, HPV-ADVISE model-country pairs in red, and the Harvard model-country pairs in blue. The grey area corresponds to the additional cases averted in the vaccinated cohort after the 100 years of routine vaccination. Health outcomes were discounted at 3% (panels A–D) and 0% (panels E–H).

Across all income groups, the threshold (i.e., maximum) cost for the second dose to remain cost-effective was low—from 0.85 (range of medians: 0.07–3.82) USD in low-income countries to 18.08 (range of medians: -3.62–85.64) USD in high-income countries if one-dose confers 30 year protection to—as few additional cancers would be averted with a longer duration of protection (≥ 30 years) or higher VE (>80%). With a higher GDP per capita, middle- and high-income countries have a higher threshold cost (Fig 5). However, if one-dose confers ≤20 years of protection, the threshold cost for the second dose to remain cost-effective is borderline at 3.24 (range of medians: 0.34–5.35) USD in low-income countries and 62.93 (range of medians: 12.63–117.45) USD in high-income countries, suggesting that duration of protection remains the main driver of uncertainty.

**Figure 5.**
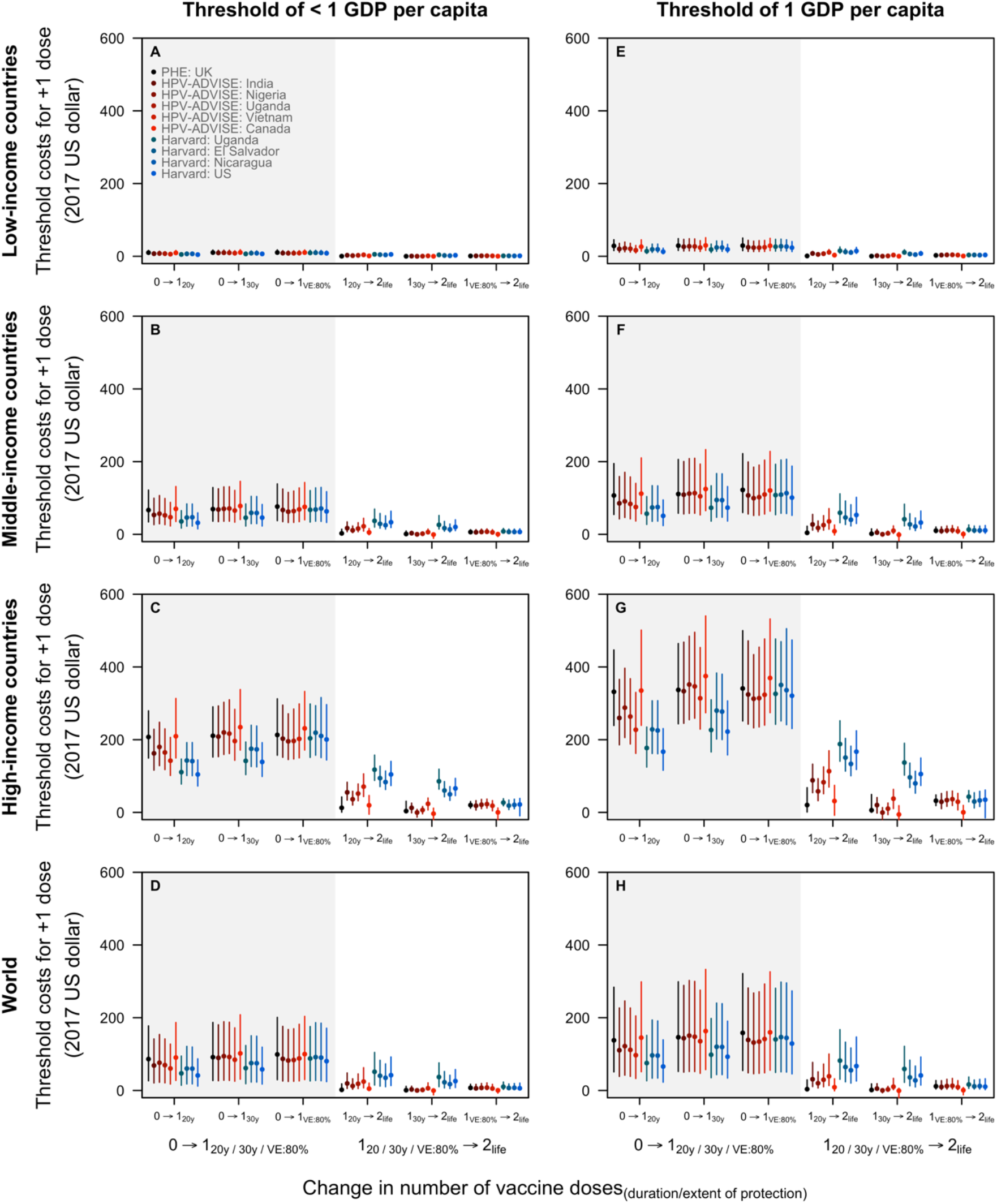
Threshold cost to pay for the first and second dose of vaccine by country income groups. The threshold cost is the maximum that could be paid for the first dose (compared to no vaccination) and second dose (compared to one dose only) for the incremental cost-effectiveness ratio to remain below the cost-effectiveness threshold. Two cost-effectiveness thresholds are presented: a lower threshold as suggested by Jit (2020) in panels A–D and country gross domestic product (GDP) per capita (in 2017 USD) in panels E–H. The lower cost-effectiveness threshold presented in panels A–D is 30–40% and 60–65% of GDP per capita in low-income and middle- to high-income countries, respectively. Cost and health outcomes were discounted at 3% and 0%, respectively.

## Discussion

In this study, three independent transmission dynamic models projected consistent results suggesting that routine one-dose HPV vaccine programmes at 80% coverage worldwide could provide a high level of population protection and be cost-effective. We considered three assumptions of the one-dose schedule: one dose of the HPV vaccine confers either 20 or 30 years of protection at full VE, or lifelong protection but at 80% VE. Across all assumptions, one-dose schedules provide large population impacts on cervical cancer, while the difference in population impact of the one-dose versus two-dose vaccination schedule is small if one dose confers ≥ 30 years of protection or lifelong protection but at 80% VE. This underscores the significant potential public health impact of the one-dose vaccination schedule if vaccine uptake is high across all countries (21).

Although trials (28–30) and post-randomisation analyses (30) suggest that the duration of protection of one dose of the vaccine is more than ten years, it is uncertain how long vaccinated individuals will remain protected and how the vaccine would wane beyond the first decade. The threshold duration of protection for a one-dose schedule to avert the majority of vaccine-preventable cancers is associated with the ages at which individuals are vaccinated and reach peak sexual activity, which varies between countries. If one dose of the vaccine confers ≤20 years of protection, giving the second dose may have a larger health impact, especially in settings where HPV transmission persists decades after vaccination. However, if one dose provides a longer duration of protection (≥ 30 years), administering the second dose will bring about few health gains at potentially high costs. Hence, decisions on offering the second dose should account for the duration of protection provided by the first dose, whether it covers the peak years of sexual activity and HPV transmission, and the costs of delivering the additional dose. The second dose becomes more cost-effective if the protection from one dose is less than 20 years, the costs of the vaccine and delivering it are lower than current reported costs, and/or the local burden of cervical cancer is high.

Our comparison of one- and two-dose vaccination schedules is motivated by several advantages of a one-dose schedule. Firstly, many LMICs have yet to implement national HPV vaccination programmes because of the challenges of delivering two vaccine doses to adolescent females (17). Compared to two-dose HPV vaccination, a one-dose HPV vaccination schedule would be cheaper and easier to implement (e.g., no follow-up of vaccinated individuals would be required), potentially enabling more LMICs to introduce HPV vaccine into national immunisation schedules (21,23). More recently, HPV vaccine implementation in LMICs has been delayed due to constraints in HPV vaccine supply (20,21). Our model-based analysis predicts that routinely vaccinating 10-year-old girls at 80% coverage in LICs could result in four times (population-adjusted) more cervical cases averted than in high-income countries. Under our one-dose assumptions, routine one-dose HPV vaccination programmes could protect up to 155 million females against cervical cancer globally over the years 2021–2120.

Secondly, the COVID-19 pandemic has disrupted several routine immunisation programmes (50–52), including HPV vaccination (21,53). Abbas and colleagues predicted that the benefits of resuming routine childhood immunisation services outweigh the risk of being infected with COVID-19 during the vaccination visits (52), reinforcing WHO’s call for all countries to continue routine immunisation services safely (54). With physical distancing measures such as school closures and national lockdowns being implemented in many countries to cope with the COVID-19 pandemic (22), health officials grapple with reconfiguring school-based HPV vaccine delivery (53–55). Compared to the two-dose vaccination schedule, a one-dose schedule would further minimise interactions between vaccinees and health workers, simplifying vaccine delivery while also decreasing SARS-CoV-2 exposure.

The lack of country-specific behavioural, virological, and clinical data in many countries limits fitting transmission dynamic models individually to most countries (56). However, in this comparative modelling study, we synthesised results from three published dynamic models based in nine countries, covering high-, middle- and low-income settings across three continents and a wide variety of epidemiological characteristics for HPV transmission and cervical cancer (14,56). Our approach provides a common framework using PRIME for population demographics, cervical cancer burden, and impact/cost-effectiveness calculations, while varying representation of HPV transmission and cervical cancer natural history across the three dynamic models. We then extrapolated the age- and time-dependent ratio of the secondary to primary impacts of vaccine strategies to other countries. While there may be considerable uncertainty around extrapolating this ratio to another country, the use of 10 model-country pairs lends confidence that we are likely to have captured the range of possible outcomes for most countries. More precise estimates would require fitting these models to additional specific countries, for which calibration data are not available (56,57).

Our model projections of vaccine impact also involve other sources of uncertainty that we did not explicitly quantify. The PRIME model uses country-specific cervical cancer burden from the Global Cancer Incidence, Mortality and Prevalence (GLOBOCAN) database (58), which may underestimate the full burden of HPV-related disease, and thus vaccine impact, in LMICs (14). In this study, we only assessed the effect of HPV vaccination on cervical cancers. If we also accounted for the vaccine impact on other HPV-related cancers, we would anticipate a greater value of HPV vaccination programmes (26,59). However, the paucity of data on the efficacy of one dose on non-cervical cancers complicates the analysis evaluating their vaccine impact. Because the health gains from the second dose are small, any minor variations in gains will amplify the variability in the number needed to vaccinate with the additional dose. Finally, we project the impact of HPV vaccination on cervical cancers over the next century. Over the past decades, we have witnessed substantial demographic (60) and behavioural changes (61,62) with extraordinary improvements in public health (63). In 2020, the COVID-19 pandemic has caused substantial disruptions to population demography (64) and sexual behaviour (65), with uncertainty around the longer-term consequences of such disruption. Moreover, over the next century, we expect to see continued advancements in pre-cancer screening and treatment services, which will further decrease cervical cancer incidence. Such uncertainties in life expectancy, population, and economic forecasts have significant implications for our predictions.

## Conclusion

Under the scenarios where a single HPV vaccine dose confers more than 30 years of protection or 80% efficacy with lifelong protection, routine one-dose HPV vaccination provides the majority of health benefits to the two-dose programme while simplifying vaccine delivery, reducing costs, and circumventing vaccine supply constraints. The second dose may be cost-effective if there is a shorter duration of protection from one dose, cheaper vaccine and vaccination delivery strategies, and high burden of cervical cancer. These results are fairly consistent when projected from three independent transmission dynamic models used in nine countries. The outcomes of our comparative modelling analysis contribute to the extensive evidence base, including emerging evidence from the single-dose HPV vaccine trials and observational studies, which would be beneficial to policymakers when they consider HPV vaccination in their populations.

## Data Availability

All analysis codes are available at https://github.com/kieshaprem/hpv-1-dose.

## Abbreviations

DALY: Disability-adjusted life year
GDP: Gross domestic product
HPV: Human papillomavirus
LMICs: Low- and middle-income countries
PHE: Public Health England
PRIME: Papillomavirus Rapid Interface for Modelling and Economics
RR: Risk ratios
UI: Uncertainty intervals
UK: United Kingdom
US: United States
USD: United States Dollar VE Vaccine efficacy
WHO: World Health Organization

## Authors’ Contributions

KP and MJ conceptualised the study, curated and analysed the data, interpreted the findings, and drafted manuscript. YHC, EB, EAB, LH, and JFL curated and analysed the data, interpreted the findings, and were major contributors in writing the manuscript. JJK and MB interpreted the findings and were major contributors in writing the manuscript. MCR, MD, SS, KA, and AP analysed the data, interpreted the findings, and reviewed the manuscript. All authors read and approved the final manuscript.

## Data availability

All analysis codes are available at https://github.com/kieshaprem/hpv-1-dose.

## Acknowledgement

We thank members of the Single-Dose HPV Vaccine Evaluation Consortium for comments and helpful discussion on this work.

