## Supplementary Materials for "Global impact and cost-effectiveness of one-dose versus two-dose human papillomavirus vaccination schedules: a comparative modelling analysis"

Supplementary Material

Kiesha Prem*, Yoon Hong Choi†, Élodie Bénard†, Emily A Burger†, Liza Hadley, Jean-François Laprise, Catherine Regan, Mélanie Drolet, Stephen Sy, Kaja Abbas, Jane J Kim, Marc Brisson, Mark Jit‡

**References** **23**

### 1 Materials and methods

#### 1.1 Model description

The three transmission dynamic models listed in Table 1—PHE, HPV-ADVISE, Harvard—were developed independently, but have several common features (1–7). The models stratify population by age, sex and sexual activity-based risk group, and screening behaviour-based risk group. They capture HPV natural history and disease, as well as HPV transmission as informed by country-specific sexual behaviour surveys.

We use a hybrid approach: first, we consider the age-specific impact that HPV vaccines may have using the results of the three independent HPV transmission dynamic models across 10 settings (1–6), and second,

†Contributed equally

Table 1: Transmission dynamic model description.

| Model configuration | PHE | HPV-ADVISE | Harvard |
|---|---|---|---|
| Countries considered | United Kingdom | India, Nigeria, Uganda, Vietnam | United States, Uganda, Nicaragua, El Salvador |
| Age cohorts | Cohorts of 10–74 years old females and males | Open, stable population from 10-years-old until death | Cohorts of 10–99 years old females and males |
| Routine vaccination | 10-year-old girls | 10-year-old girls | 9-year-old girls |
| First-year catch-up campaign | 11–14-year-old girls | 11–14-year-old girls | 10–14-year-old girls |
| Simulations | Probabilistic sensitivity analysis using second-order simulation methods | | |
| Cancer outcomes | Age-specific cervical cancer incidence | | |

extrapolate these effects to the remaining countries in the world using data on population demographics and cervical cancer burden synthesised in a static model Papillomavirus Rapid Interface for Modelling and Economics (PRIME) model (8,9).

#### 1.2 The HPV-ADVISE models

The HPV-ADVISE (2,3) and HPV-ADVISE LMIC (10) models are individual-based, transmission-dynamic models of multi-type HPV infection–with 18 HPV types modelled separately, including vaccine-preventable types 6/11/16/18/31/33/45/52/58—and diseases. Designed to examine HPV vaccination policy questions in low- and middle-income countries (LMICs) settings, HPV-ADVISE LMIC has the same underlying structure for all LMICs and the same basic model structure as HPV-ADVISE for high-income countries (HICs) but modified to capture differences in sexual behaviour between LMICs and HICs. More details can be found in the Technical Appendices: http://www.marc-brisson.net/HPVadvise.pdf and http://www.marc-brisson.net/HPVadvise-LMIC.pdf. HPV-ADVISE and HPV-ADVISE LMIC reproduce demographic characteristics, sexual behaviour and transmission of HPV, the natural history of HPV-associated diseases (HPV infection, natural immunity, three grades of cervical lesions, and three cervical cancer stages), screening and treatment. Transmission is gender- and age-specific, and depends on sexual behaviour (e.g., mixing patterns) and HPV biology and natural history (e.g., probability of transmission and natural immunity). For each country, we identified 50 parameter sets that simultaneously fit highly stratified country-specific sexual behaviour and HPV epidemiological data obtained from published articles, specific studies, and international population-based datasets (HPV-ADVISE LMIC Technical Appendix Table A1). These 50 parameter sets represent uncertainty in model parameters and variability in sexual behaviour and HPV epidemiology within the modelled country. They were used to generate age-specific incidence of cervical cancer over time for each vaccination scenario investigated.

#### 1.3 The Harvard model

The Harvard model (5,6) uses a multi-model approach to project the population health consequences of alternative cervical cancer scenarios over time. The multi-modelling approach involves two components: (1) Harvard-HPV, a dynamic, compartmental model of natural history that simulates the potential health outcomes of nine HPV infection genotypes and HPV sexual transmission between males and females; and (2) Harvard-CC, a static, individual-based model of HPV-induced cervical cancer. We simulated HPV transmission in Harvard-HPV as a function of partnership acquisition and dissolution, by sex, age, and sexual activity level. HPV infection can be transmitted, depending on the number of new partners, partner infection status, probabilities of HPV transmission given contact with an infected partner, and duration of the partnership. Individuals develop type-specific natural immunity when they clear an HPV infection, which reduces their susceptibility to future same-type infection. Harvard-HPV projects reductions in HPV incidence by genotype over time associated with each control strategy; these reductions served as inputs into Harvard-CC. Using Harvard-CC, we project cervical cancer incidence by age over time for each scenario as a series of transitions through health states that describe true underlying health, including HPV infection, precancerous states, and invasive cancer. Women who develop cervical cancer may be detected symptomatically or progress to a more severe cancer stage. The model is adapted to different epidemiological settings—the United States, Uganda, El Salvador and Nicaragua—by fitting or calibrating the model using the best available country-specific data, e.g., HPV prevalence, HPV type distribution in CIN 2, CIN3 and cervical cancer. More details about the model can be found at: https://cisnet.cancer.gov/cervical/profileshtml.

#### 1.4 The Public Health England (PHE) model

The Public Health England model(1) is a parameterised family of dynamic transmission models of heterosexual HPV transmission and HPV-related diseases—cervical dysplasia, cervical cancer and anogenital warts—in the United Kingdom. At equilibrium, the model population consists of 49 million people. They are divided into birth cohorts between 10 and 74 years old, with an equal split between males and females. Age- and sex-specific mortality rates were obtained from the Office for National Statistics of the UK. The model population is closed, and it assumes no immigration or emigration. This series of transmission models represent different parameters about HPV biology and epidemiology, including sexual behaviour, natural immunity, vaccine characteristics, disease progression and screening accuracy with thousands of combinations of assumptions on the parameters generated and then fitted to prevalence data. Each scenario or combination of assumptions was then fitted to epidemiological data and the best-fitting scenarios used to predict the impact of vaccination.

#### 1.5 Papillomavirus Rapid Interface for Modelling and Economics (PRIME) model

The Papillomavirus Rapid Interface for Modelling and Economics (PRIME) model(8,9) is a WHO-supported model to estimate the health impact—cervical cancer cases, deaths, or disability-adjusted life-years averted—and cost-effectiveness of HPV vaccination strategies among adolescent girls. As a static model, PRIME does not consider herd effects and cross-protection against non-vaccine HPV types and thus provides conservative estimates of the vaccine impact. However, it adjusts for lower vaccine protection in vaccinated individuals who have sexually debuted before vaccination. The updated PRIME(9) uses population demography of the United Nations World Population Prospects 2019 revision(11) to incorporate population ageing. In the updated model, cervical cancer burden was updated from the International Agency for Research on Cancer estimates for Global Cancer Incidence, Mortality and Prevalence (GLOBOCAN) 2012(12) database to GLOBOCAN 2018 database(13), and disability weights were updated from estimates of the Global Burden of Disease 2001 study(14) to estimates of the 2017 study(15). PRIME can also estimate the health impact of bivalent or quadrivalent and nonavalent vaccination programmes. Disability-adjusted life year (DALY) was estimated as the sum of years of life lost due to premature mortality and years of life lost due to time lived in states of less than full health due to disability (more details in Appendix Tables A2.1. and Tables A2.2. in Abbas and colleagues (9)). The disability weights for different phases (diagnosis and primary therapy, controlled, metastatic, terminal) of cervical cancer from the Global Burden of Disease 2017 study were used to estimate the years of life lost due to disability. The country-specific life tables were used to estimate the years of life lost due to premature mortality. The treatment cost, including facility, staff, medical device and pharmaceutical costs, for cancers detected at each stage in 14 WHO-CHOICE regions was obtained from a WHO-CHOICE study (16).

#### 1.6 Vaccination strategies

We model routine annual vaccination with the 9-valent vaccine in 10-year-old girls to begin in 2021 and run uninterrupted until 2120. We also include a catch-up of girls aged 11–14-year-old in the first year of the programme. Vaccine coverage was assumed to be 80%. We measure and compare population-level impact (e.g., cervical cancers averted, number of females needed to be vaccinated, threshold costs of the first and second dose of the vaccine) for three vaccine strategies: no HPV vaccination; a one-dose HPV vaccination schedule in which we assume that one dose of the vaccine gives either a shorter duration of protection (20 or 30 years) or lower vaccine efficacy (e.g., 80%) compared to two doses; and a two-dose HPV vaccination schedule in which two doses of the vaccine would provide lifetime protection. Although the Harvard, HPV-ADVISE and PHE models incorporated vaccine efficacy differently—vaccine 'degree' for the Harvard model and vaccine 'take' for HPV-ADVISE and PHE—the models achieved an 80% cumulative reduction in vaccine-type HPV infections for a vaccinated cohort at year five.

Table 2: Vaccination strategies.

| **Parameters** | **Description** |
|---|---|
| Vaccine type | HPV types prevented by vaccination: all high-risk HPV types in the 9-valent vaccine (16, 18, 31, 33, 45, 52, and 58) |
| Age of vaccination | Age of routine vaccination and ages covered by multi-age cohort (MAC) in first year: Routine 10-year-old girls including first year MAC for 11–14 years old girls |
| Coverage of vaccination | Proportion of girls in targeted age groups who are vaccinated: 80% |
| Years of vaccination | Year 1–101 (or calendar years 2021–2120); year 0 is pre-vaccination |
| Gender | Females |
| Vaccination scenarios | **Scenario 0:** no vaccination<br>**Scenario 1:** lifetime protection, vaccine efficacy (VE): 100%, i.e., similar to the current two-dose assumptions<br>**Scenario 2:** one-dose offers 20 years protection, VE: 100%<br>**Scenario 3:** one-dose offers 30 years protection, VE: 100%<br>**Scenario 4:** one-dose offers lifetime protection, VE: 80% protection against persistent infection at 5-year time point. |

#### 1.7 World Bank income groups

The World Bank assigns countries in the world to three income groups—low, middle, and high-income countries—based on gross national income per capita in current USD of the previous year. Table 3 lists the 192 countries (as ISO 3 country codes) included in the study by their income group.

#### 1.8 Population projection

In this analysis, we model health outcomes in females born in the years 2011–2110. We accrue all health benefits of HPV vaccination up to the end of the routine vaccination programme (i.e. the year 2120) or age 100 of all vaccinated cohorts (i.e., up to the year 2210). The United Nations Population Division projects the population for all countries and areas of the world up to the year 2100 (17). We use these projections up to 2100, and we then project the population for all countries for the years 2101–2210. We ran a demographic model to age the population in 2100, depleting it by deaths and replenishing it with births.

**Fertility rate model:** For the demographic model, we used projected five-year age-specific fertility rates for the 192 countries over the period 2095 to 2100, obtained from the United Nations Department of Economic and Social Affairs (17), as a baseline. The age-specific fertility rates were then held fixed in the model to the year 2210.

Table 3: World Bank income group.

| Income group | Country code |
|---|---|
| Low-income | AFG, BDI, BFA, CAF, COD, ERI, ETH, GIN, GMB, GNB, HTI, LBR, MDG, MLI, MOZ, MWI, NER, PRK, RWA, SDN, SLE, SOM, SSD, SYR, TCD, TGO, TJK, UGA, YEM |
| Middle-income | AGO, ALB, ARG, ARM, AZE, BEN, BGD, BGR, BIH, BLR, BLZ, BOL, BRA, BTN, BWA, CHN, CIV, CMR, COG, COL, COM, CPV, CRI, CUB, DJI, DOM, DZA, ECU, EGY, FJI, FSM, GAB, GEO, GHA, GNQ, GRD, GTM, GUY, HND, IDN, IND, IRN, IRQ, JAM, JOR, KAZ, KEN, KGZ, KHM, KIR, LAO, LBN, LBY, LCA, LKA, LSO, MAR, MDA, MDV, MEX, MKD, MMR, MNE, MNG, MRT, MYS, NAM, NGA, NIC, NPL, PAK, PER, PHL, PNG, PRY, PSE, RUS, SEN, SLB, SLV, SRB, STP, SUR, SWZ, THA, TKM, TLS, TON, TUN, TUR, TZA, UKR, UZB, VCT, VEN, VNM, VUT, WSM, ZAF, ZMB, ZWE |
| High-income | ARE, ATG, AUS, AUT, BEL, BHR, BHS, BRB, BRN, CAN, CHE, CHL, CYP, CZE, DEU, DNK, ESP, EST, FIN, FRA, GBR, GRC, GUM, HRV, HUN, IRL, ISL, ISR, ITA, JPN, KOR, KWT, LTU, LUX, LVA, MLT, MUS, NCL, NLD, NOR, NZL, OMN, PAN, POL, PRI, PRT, PYF, QAT, ROU, SAU, SGP, SVK, SVN, SWE, SYC, TTO, URY, USA |

Because much about the evolution of fertility rates after reaching replacement levels remains unknown and may vary due to cultural differences between countries, we did not seek to extrapolate the fertility trend beyond the latest available projections. Instead, we kept the age-specific fertility rate at 2095 constant until 2210, possibly overestimating fertility as a result. Using the UN's projected sex ratio at birth over the period 2095 to 2100, we distributed the expected births to males and females. We denote $b_m$ to be the proportion of births that are male in country $c$.

Letting $\phi_a^c$ be the fertility rate for women aged $a$ in country $c$, and $F_a^c(t)$ be the number of women aged $a$ in country $c$ in year $t$, we derived the expected number of births in each country and year to be

$$E^c(t) = \sum_a \phi_a^c F_a^c(t).$$

The number of children surviving to age one was derived by separately calculating the expected number of male births

$$B^c(t) = E^c(t) b_m$$

and female births

$$G^c(t) = E^c(t)(1 - b_m).$$

**Mortality rate model:** Projected annual, sex-specific, five-year mortality rates were available for the years 2095 to 2100 from the life tables obtained from the United Nations Department of Economic and Social Affairs. The life tables by sex were up to age 100, and they provide projections of the mortality experience of a hypothetical group of infants born at the same time and subject throughout their lifetime to the specific mortality rates of the years 2095–2100. These were also available for the 192 countries. The mortality rate was assumed constant over five-year intervals of age (with those below 1-year-old having their own mortality group, and those above 100-years-old aggregated in one age group). Using the country-specific life tables, we aged the population forward in time and depleting it by deaths estimated using the age- and country-specific mortality rates.

#### 1.9 HPV-FRAME reporting standard checklist

The checklists presented in Tables 4–6 include the reporting standards from HPV-FRAME (18).

Table 4: Inputs: HPV-FRAME reporting standard checklist.

| Domain | Input | Reported by age? (Y/N) | Report by sex (F-only, M-only or both)? | Comments |
|---|---|---|---|---|
| **Core reporting standards** | | | | |
| CRS | Target population for intervention | Y | F-only | HPV vaccination of girls aged 10 years; single year of catch-up aged 11–14 years. |
| CRS | Sexual behaviour | Y | Y | Reported by age, sex and risk group[1−7] |
| CRS | Cohort examined for evaluation/time horizon | Y | F-only | 101 year time horizon (2020–2120) |
| CRS | Quality of life assumptions | N/A | N/A | Reported outcomes were cancer cases, deaths, the number needed to vaccinate and threshold costs. |
| CRS | Calibration | Y | Y | HPV-ADVISE and Harvard models reproduce Globocan 2018 incidence at a country level. The models were calibrated to sexual behaviour, HPV prevalence and cervical cancer incidence[1−7] |
| CRS | Validation | Y | F-only | Reported in [1−7] |
| CRS | Costs | N | F-only | Costs of treatment and vaccine reported in [8,9] |

Table 5: Inputs: HPV-FRAME reporting standard checklist. (Continued)

| Domain | Input | Reported by age? (Y/N) | Report by sex (F-only, M-only or both)? | Comments |
|---|---|---|---|---|
| **Reporting standard for HPV vaccination in adolescent females** | | | | |
| 1 | Vaccine uptake | Y | F-only | Described in Methods of the paper |
| 1 | Vaccine efficacy | Y | F-only | Assumed invariant by sex and age |
| 1 | Vaccine duration and waning | Y | F-only | Assumed invariant by sex and age |
| 1 | Vaccine and delivery costs | Y | Y | Vaccine and delivery costs reported in [8,9] |
| 1 | Pre-vaccination disease burden (including PAFs) | Y | F-only | Reported in [1–7] |
| 1 | Heterogeneity in sexual behaviour | Y | F-only | Reported in [1–7] |
| 1 | Duration of natural immunity | Y | F-only | Reported in [1–7] |
| **Reporting standard for HPV vaccination using alternative dose schedules** | | | | |
| 7 | Vaccine efficacy/waning (by dose, type) | Y | F-only | Assumed invariant by sex and age. We modelled full protection and 80% vaccine efficacy against HPV 16/18/31/33/45/52/58 (for 1-, 2-dose) |
| 7 | Timing between doses (for 2-dose) | N/A | N/A | Six months for the 2-dose regimens only given to girls aged 10 years in modelled scenarios. |
| 7 | Vaccine cross-protection (by dose, type) | Y implicitly | F-only | We modelled vaccination with a 9-valent vaccine where we assumed that vaccine efficacy is either 80% or 100% for HPV 16/18/31/33/45/52/58. |
| 7 | Cost per dose/per vaccinated individual | N | N/A | Cost per dose assumed constant by age within each country. |

Table 6: Outputs: HPV-FRAME reporting standard checklist.

| Domain | Output | Reported by age? (Y/N) | Report by sex (F-only, M-only or both)? | Comments |
|---|---|---|---|---|
| **Core reporting standards** | | | | |
| CRS | Cancer incidence, mortality, life years, QALYs/DALYs (as appropriate) | N | F-only | Reported outcomes were cancer cases and deaths reported combined across all ages as a total over the lifetime of females born 2011–2110 (presented in Fig 2 and the Results of the paper). |
| CRS | HPV prevalence, pre-intervention CIN2 detected | N | N | We did not report this level of detail as our study focuses on the impact of cancer occurrence and deaths. Impact of interventions on HPV prevalence and CIN2 was thus not a focus of the paper. |
| CRS | Sensitivity analysis on key inputs | Y implicitly | F-only | In this study, we compared three models with different structural and parameterisation assumptions; hence sensitivity analysis is built into the reported ranges of results between models. Additional details of the sensitivity analysis can be found in the Methods of the paper. |
| CRS | Incremental cost-effectiveness ratios and costs saved | N | F-only | Threshold costs per dose were reported. |
| **Reporting standard for HPV vaccination in adolescent females** | | | | |
| 1 | Absolute reductions in HPV infections, cervical and other HPV- related cancers and/or warts, post-vaccination | N | F-only | Reported outcomes were cervical cancer cases and deaths, not reductions in HPV infections, other HPV-related cancers or warts. Theoutcomes were combined across all ages. |
| 1 | Absolute reductions in CIN2+ post-vaccination | N | N | This paper only focuses on the reduction of cervical cancer cases, deaths and DALYs post-vaccination. |
| 1 | Absolute reductions in invasive cancer post-vaccination | N | N | Reported outcomes were cancer cases and deaths combined across all ages or as a total over the lifetime of females born 2011–2110 (year by year : Fig 2) and for females over the vaccination period 2021–2120 (year by year: Fig 2). |
| **Reporting standard for HPV vaccination using alternative dose schedules** | | | | |
| 7 | Threshold cost per dose | N | F-only | Threshold costs per dose were combined across all ages or as a total over the lifetime of females born 2011–2110. |

### 2 Results

#### 2.1 Cervical cancers cases and deaths averted

Figure 1 presents the cumulative cervical cancers cases that could have been averted by routine one-dose HPV vaccination in 192 countries over the years 2021–2120. Figure 2 shows the cervical cancers deaths that could have been prevented by routine one-dose HPV vaccination in 192 countries over the years 2021–2120. Cancer cases and deaths averted (health outcomes) are discounted at 0%. Only cervical cancer caused by HPV 16, 18, 31, 33, 45, 52 and 58, which could be averted by the 9-valent HPV vaccine, were considered. The lines represent the median projections of the 10 model-country settings: the PHE model in black, HPV-ADVISE models in red, and the Harvards models in blue. The grey area corresponds to the additional cases or deaths averted in the vaccinated cohorts after the 100 years of routine vaccination.

#### 2.2 Cervical cancer cases and deaths: discounting health outcomes at 3%

Figures 3 and 4 present the cervical cancers cases and deaths that could have been averted by routine one-dose HPV vaccination in 192 countries over the years 2021–2120. Cancer cases or deaths averted (health outcomes) are discounted at 3%. Only cervical cancer caused by HPV 16, 18, 31, 33, 45, 52 and 58, which could be averted by the 9-valent HPV vaccine, were considered. Figure 5 shows the cumulative cervical cancers cases that could have been averted by routine one-dose HPV vaccination in 192 countries over the years 2021–2120 when health outcomes are discounted at 3%.

Figures 6 and 7 present the proportion of cervical cancer cases and deaths averted by routine two-dose HPV vaccination programmes with a perfect vaccine (i.e., 100% vaccine efficacy) conferring lifelong protection that may still occur under a routine one-dose schedule. The median percentage (intervals: 10–90th percentile) of cancers not averted by a one-dose schedule compared to a two-dose program of the 10 model-country settings: the PHE model in black, HPV-ADVISE model-country pairs in red, and the Harvard model-country pairs in blue.

When we discount health benefits, the model predicts that routine vaccination would prevent fewer cancers and deaths, and more girls need to be vaccinated to avert one cervical cancer case (Figure 4 in main paper).

When we investigated the impact of a one-dose vaccination schedule with a 2-valent vaccine, the model predicts many cancers can still be averted by routine vaccination with one dose of the 2-valent vaccine (Figure 8). In 192 countries over the years 2021–2120, the models projected that routine annual vaccination of 10-year-old girls (plus a one-year catch-up campaign of girls aged 11–14 years) with one dose of the 2-valent HPV vaccine at 80% coverage would avert 50.6 million (80%UI 49–52.7) and 52.4 million (80%UI 50.6–56.8) cervical cancer cases should one dose of the vaccine confer 20 and 30 years of protection, respectively (Figure 8). Under a scenario of one dose of the 2-valent vaccine providing lifelong protection at 80% initial VE, the models predicted that 53.5 million (80%UI 51.7–57.4) cervical cancer cases would be prevented (Figure 8).

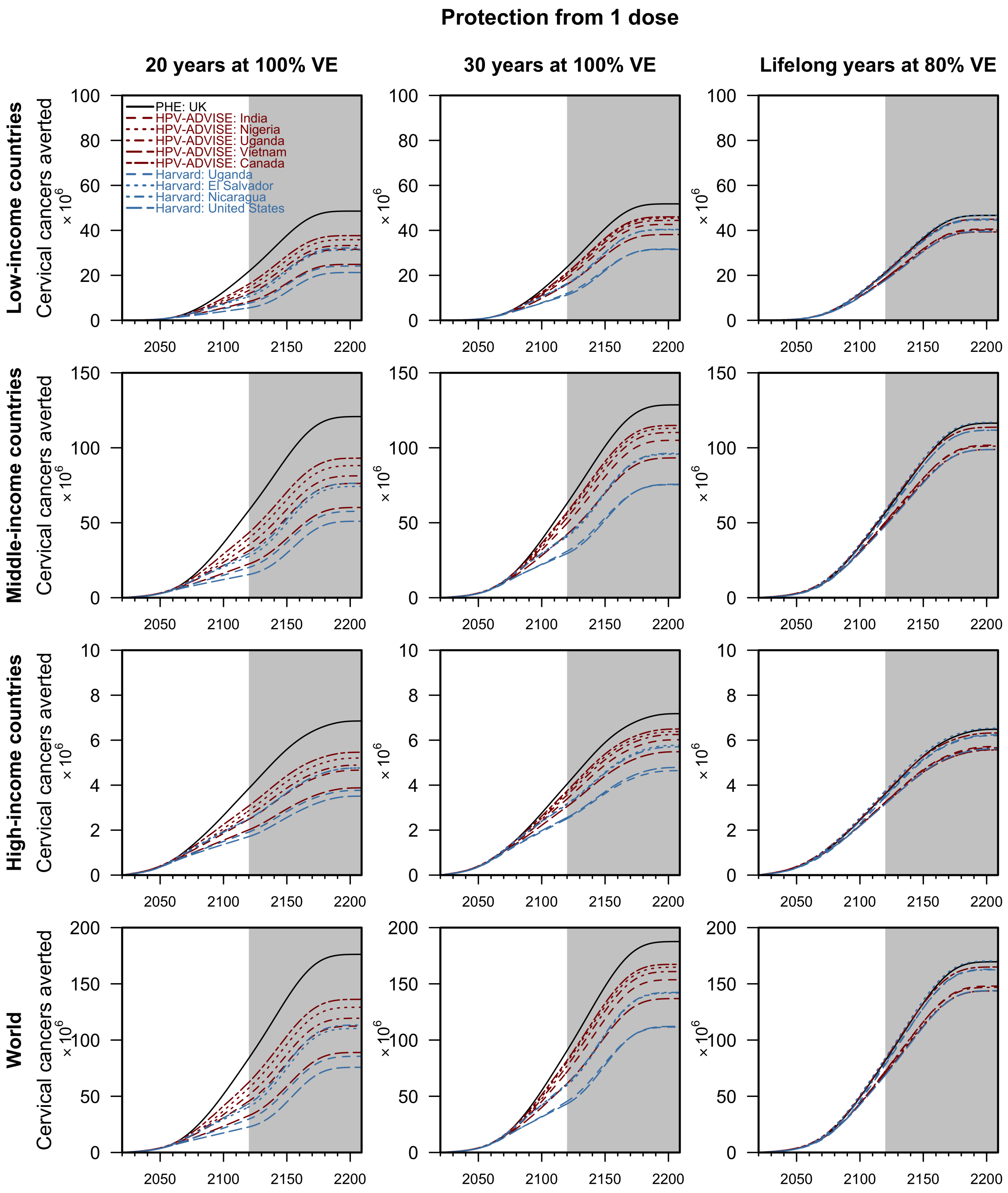

Figure 1: Cumulative cervical cancers averted by routine one-dose HPV vaccination by income groups, no discounting.

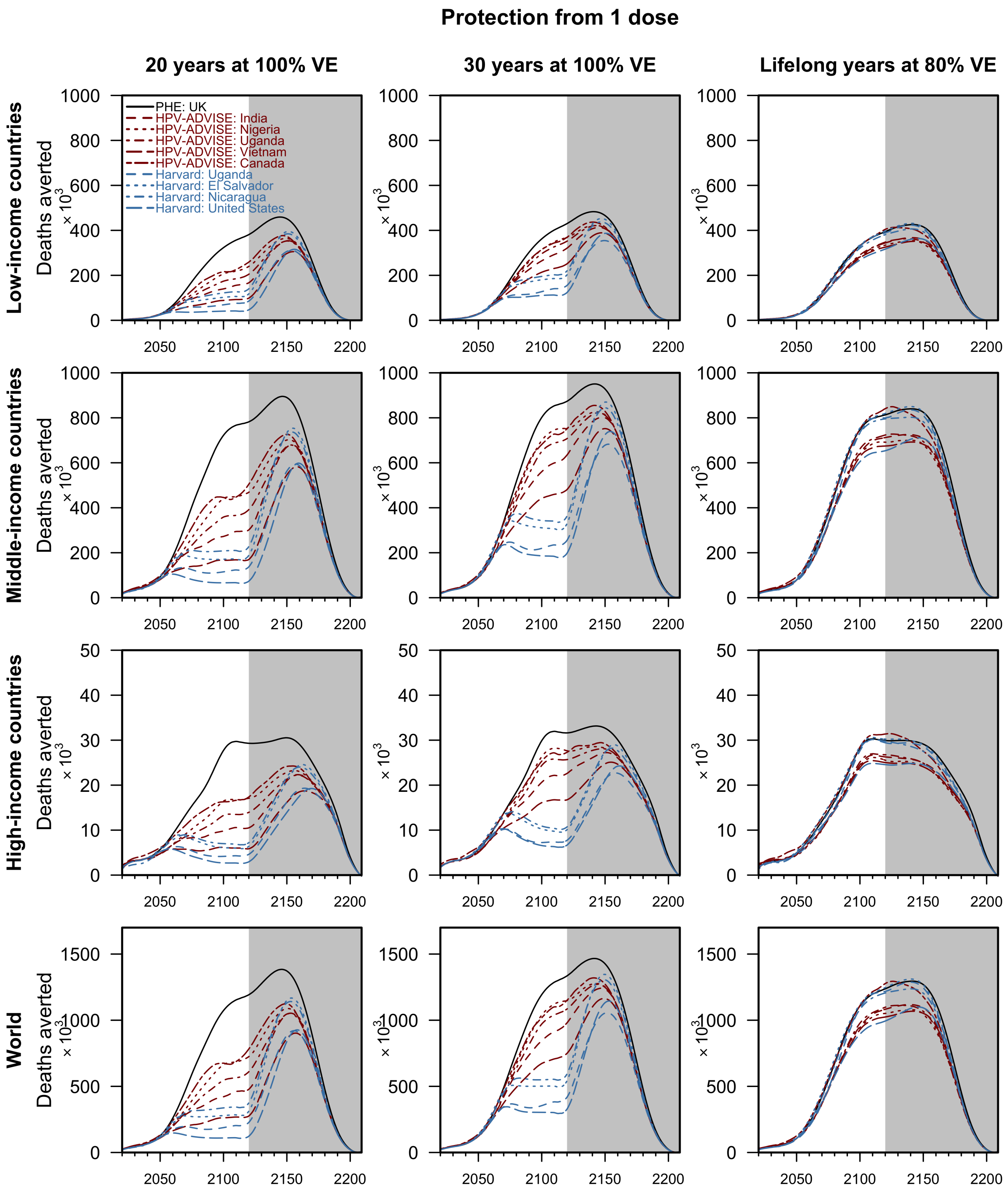

Figure 2: Cervical cancer deaths averted by routine one-dose HPV vaccination by income groups, no discounting.

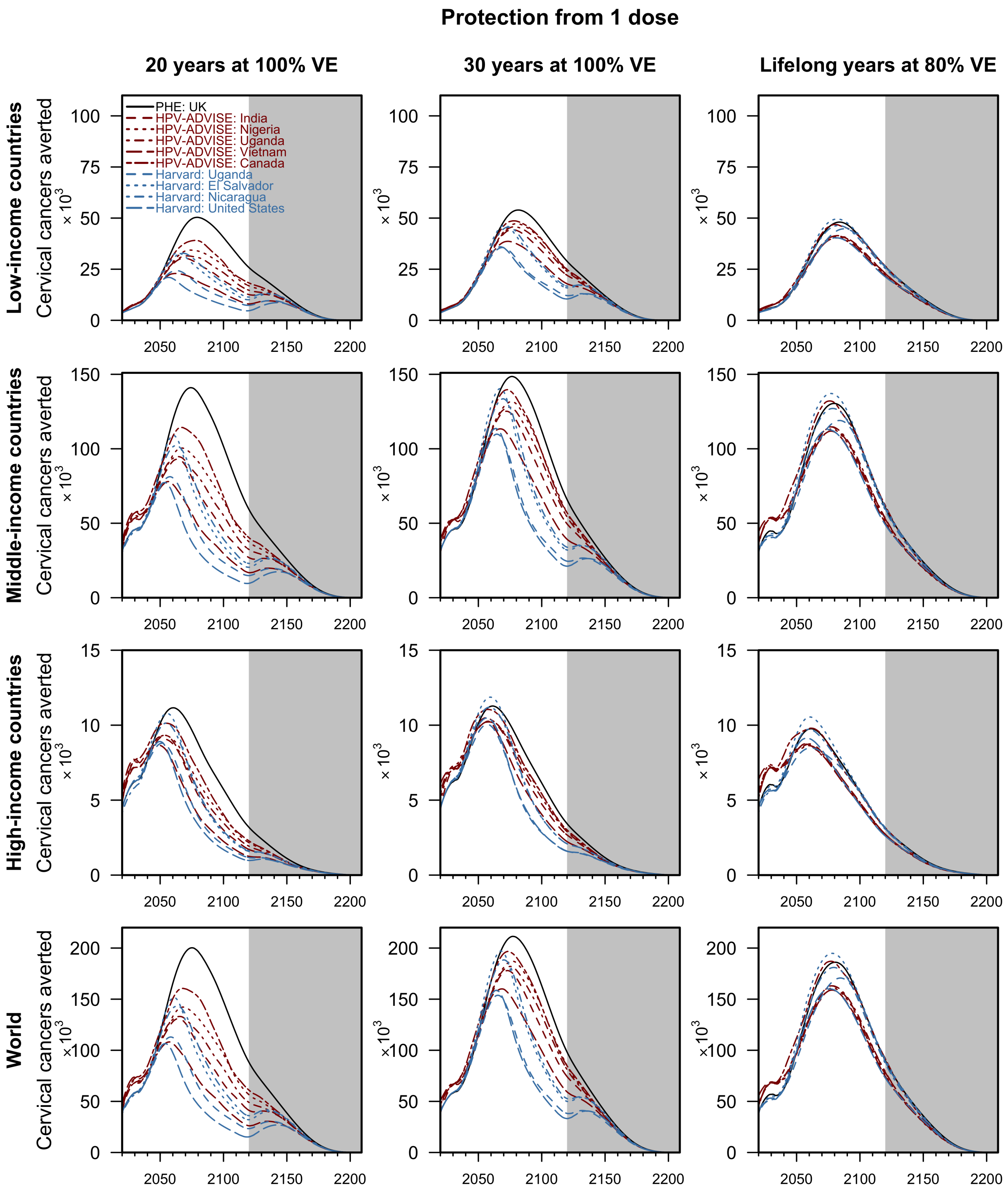

Figure 3: Cervical cancers averted by routine one-dose HPV vaccination by income groups, discounted.

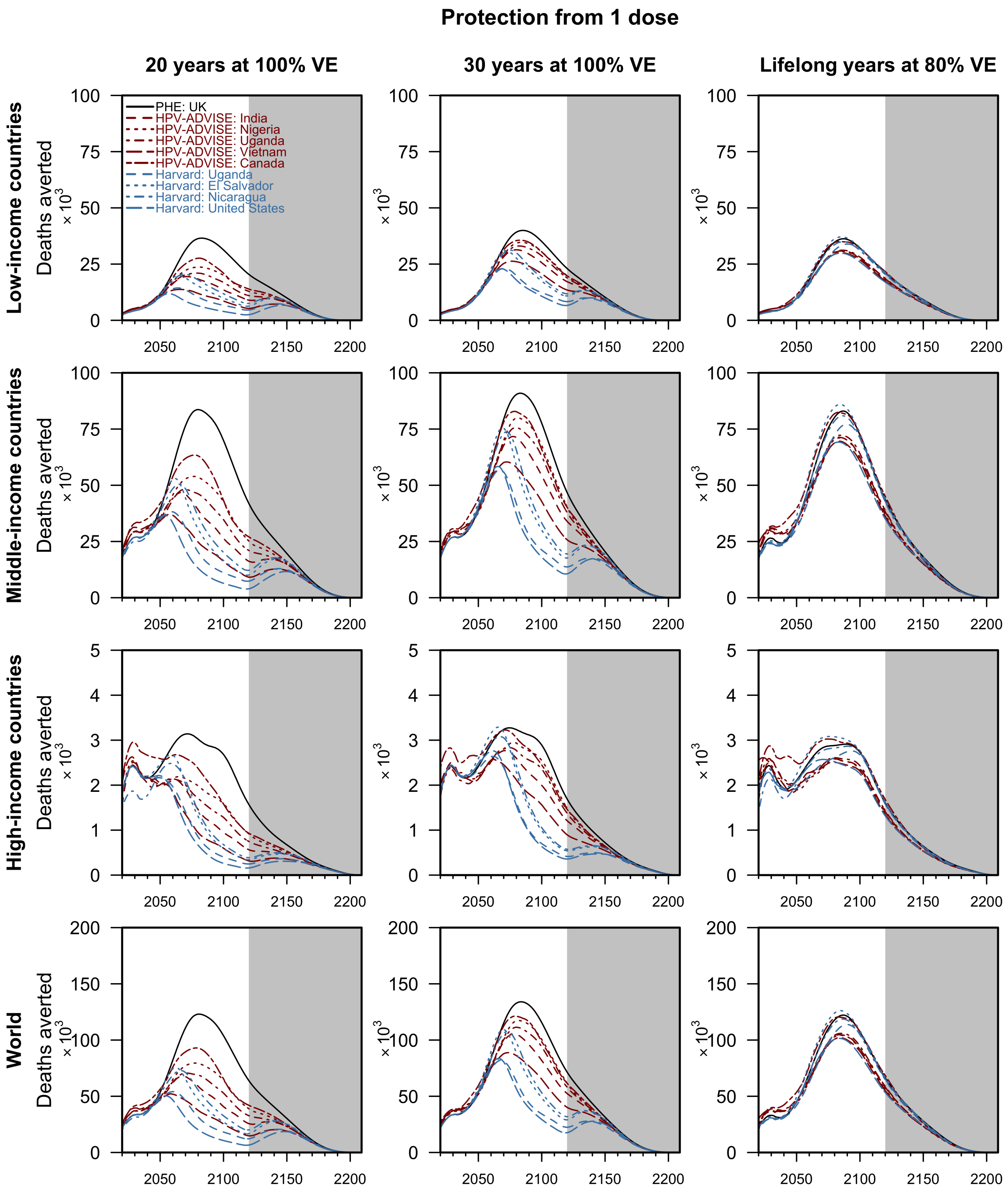

Figure 4: Cervical cancer deaths averted by routine one-dose HPV vaccination by income groups, discounted.

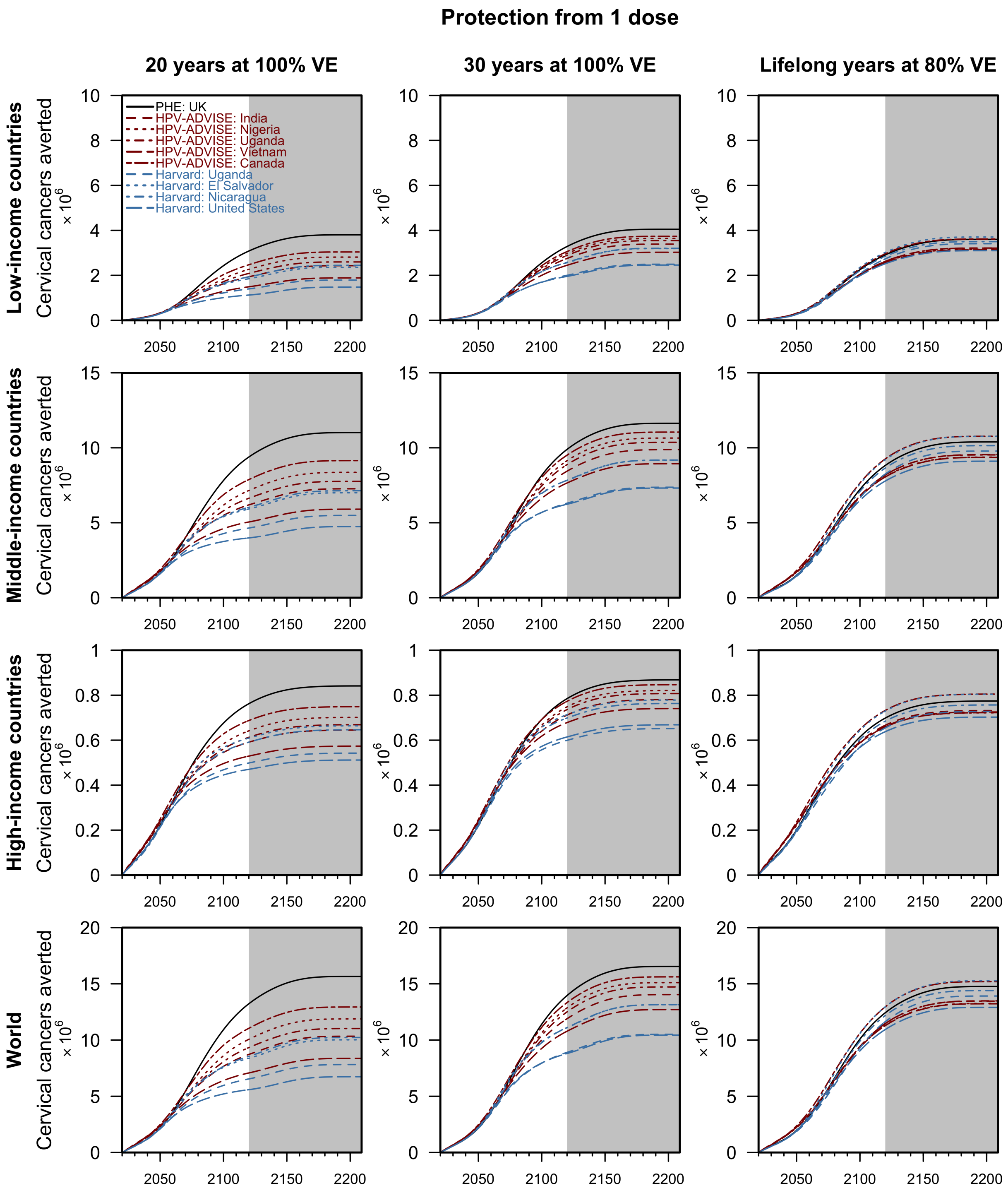

Figure 5: Cumulative cervical cancers averted by routine one-dose HPV vaccination by income groups, discounted.

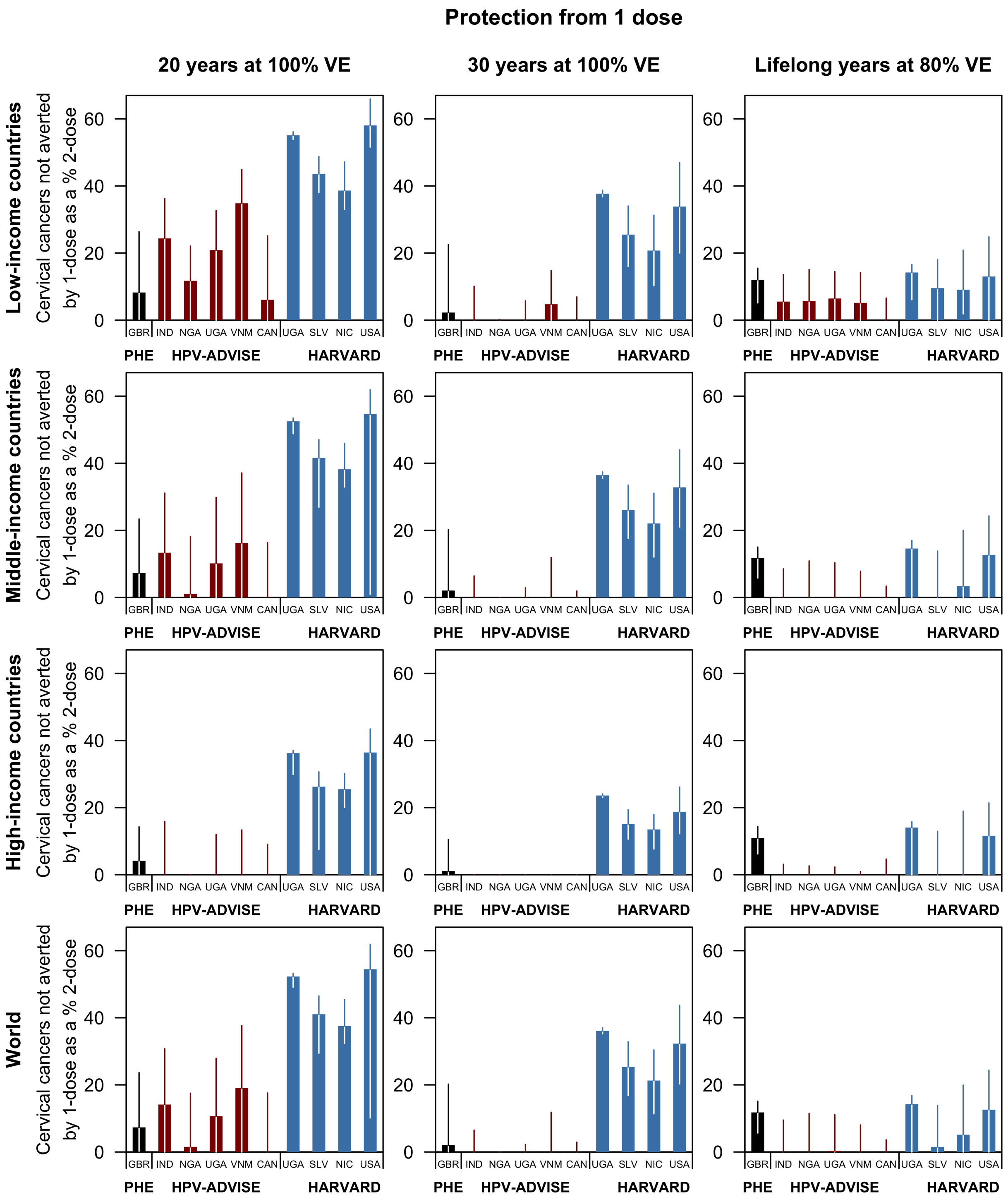

Figure 6: Proportion of cervical cancers not averted by 1-dose compared to a perfect vaccine, discounted.

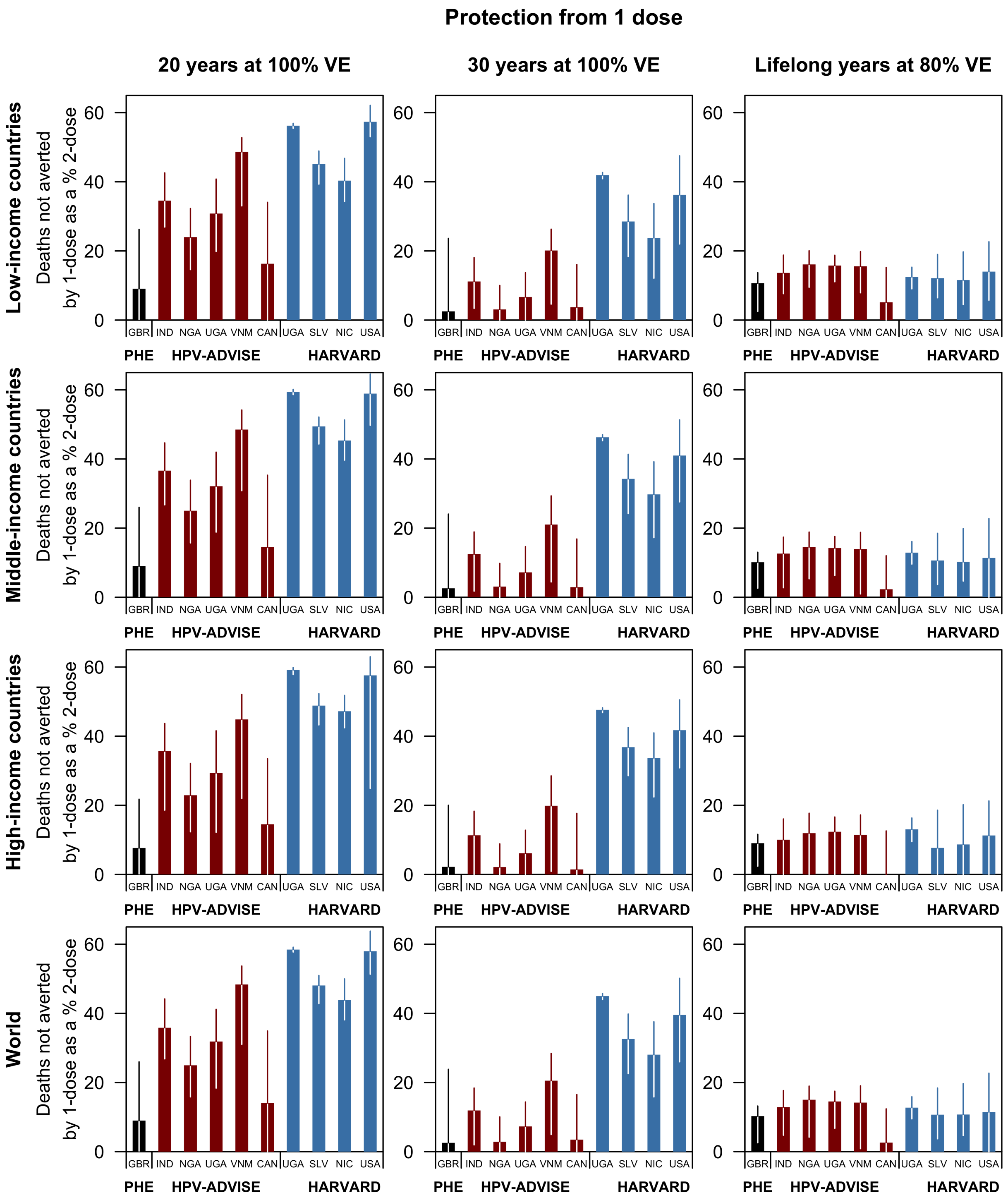

Figure 7: Proportion of cervical cancer deaths not averted by 1-dose compared to a perfect vaccine.

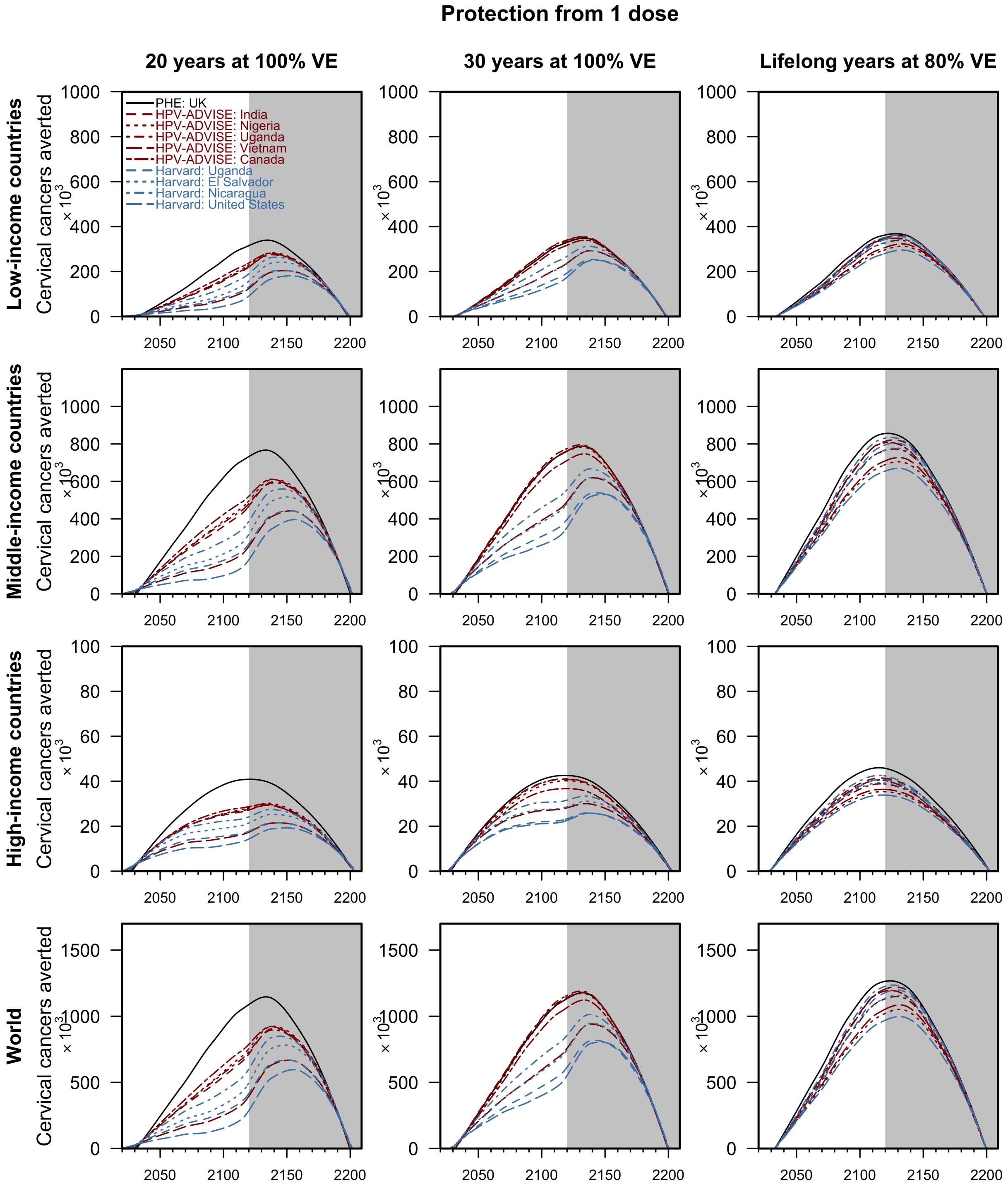

Figure 8: Cervical cancers averted by routine one-dose HPV vaccination by income groups with a 2-valent vaccine.

#### 2.3 Threshold costs

The threshold cost is the maximum that could be paid for the first dose (compared to no vaccination) and second dose (compared to one dose only) for the incremental cost-effectiveness ratio to remain below the cost-effectiveness threshold. Two cost-effectiveness thresholds are presented (20): country gross domestic product (GDP) per capita (in 2017 USD) costs in panels A–D and a lower threshold as suggested by Jit (2020)(19). The lower cost-effectiveness threshold considered is 30–40% and 60–65% of GDP per capita in low-income and middle- to high-income countries, respectively. Both health outcomes and costs are discounted at 0% and 3%. We used the GDP per capita estimates by the World Bank.

Figure 9 presents the threshold cost for the first dose (compared to no vaccination) and second dose (compared to one dose only) under two cost-effectiveness thresholds: country gross domestic product (GDP) per capita (in 2017 USD) costs in panels A–D and a lower threshold as suggested by Jit (2020)(19). The lower cost-effectiveness threshold presented in panels E–H is 30–40% and 60–65% of GDP per capita in low-income and middle- to high-income countries, respectively. Both cost and health outcomes are discounted at 3%. In Figure 10, both cost and health outcomes are not discounted.

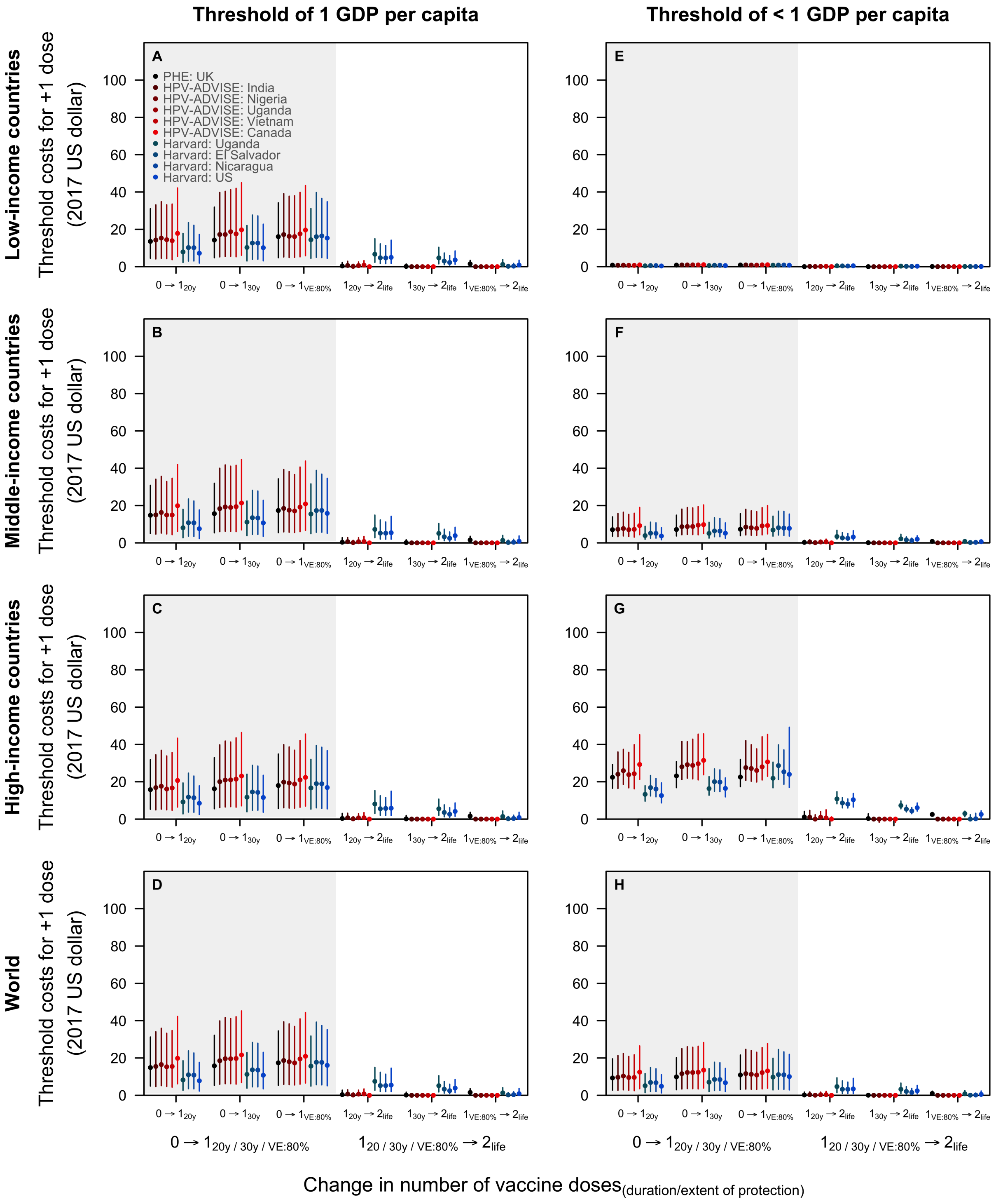

Figure 9: Threshold cost to pay for the first and second dose of vaccine, discounting on health outcomes and costs.

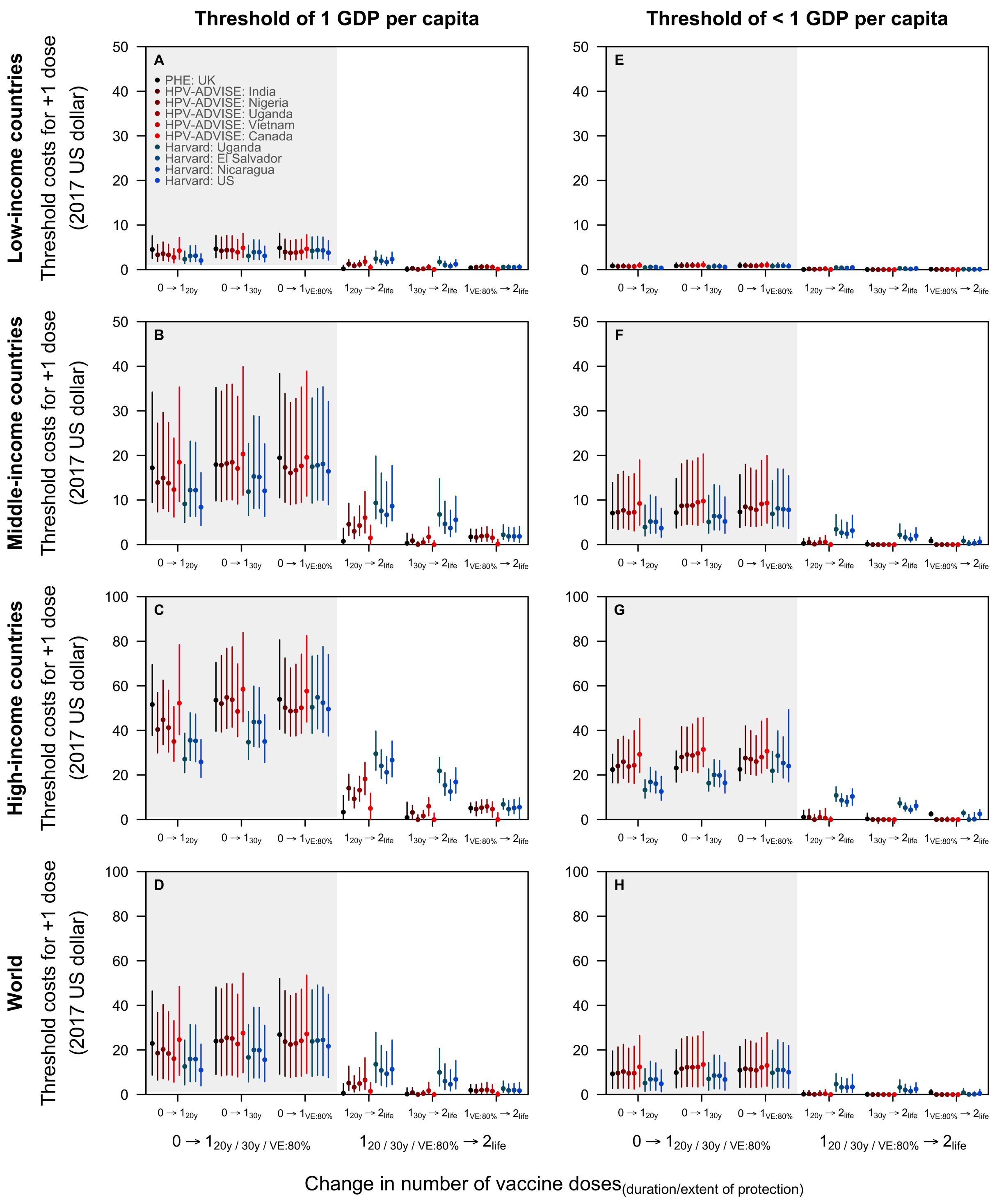

Figure 10: Threshold cost to pay for the first and second dose of vaccine, no discounting.

#### 2.4 Number needed to vaccinate

The time horizon of the analysis is from 2021 to 2120. However, we accrue all health benefits of vaccination up to the end of the routine vaccination programme (i.e. the year 2120, in the figure below) or age 100 of all vaccinated cohorts. Over the years 2021 to 2120, Figure 11 shows the number of girls needed to be vaccinated with the first and second dose to avert one additional cervical cancer case by income group when health outcomes are discounted at 3% (panels A–D) and 0% (panels E–H). The lines represent the median projections of the nine models: the PHE model in black, HPV-ADVISE models in red, and the Harvards models in blue. Health outcomes are discounted at 3% (panels A–D) and 0% (panels E–H). In the main paper, we present the results when we accrue health benefits of vaccination up to age 100 of all vaccinated cohorts.

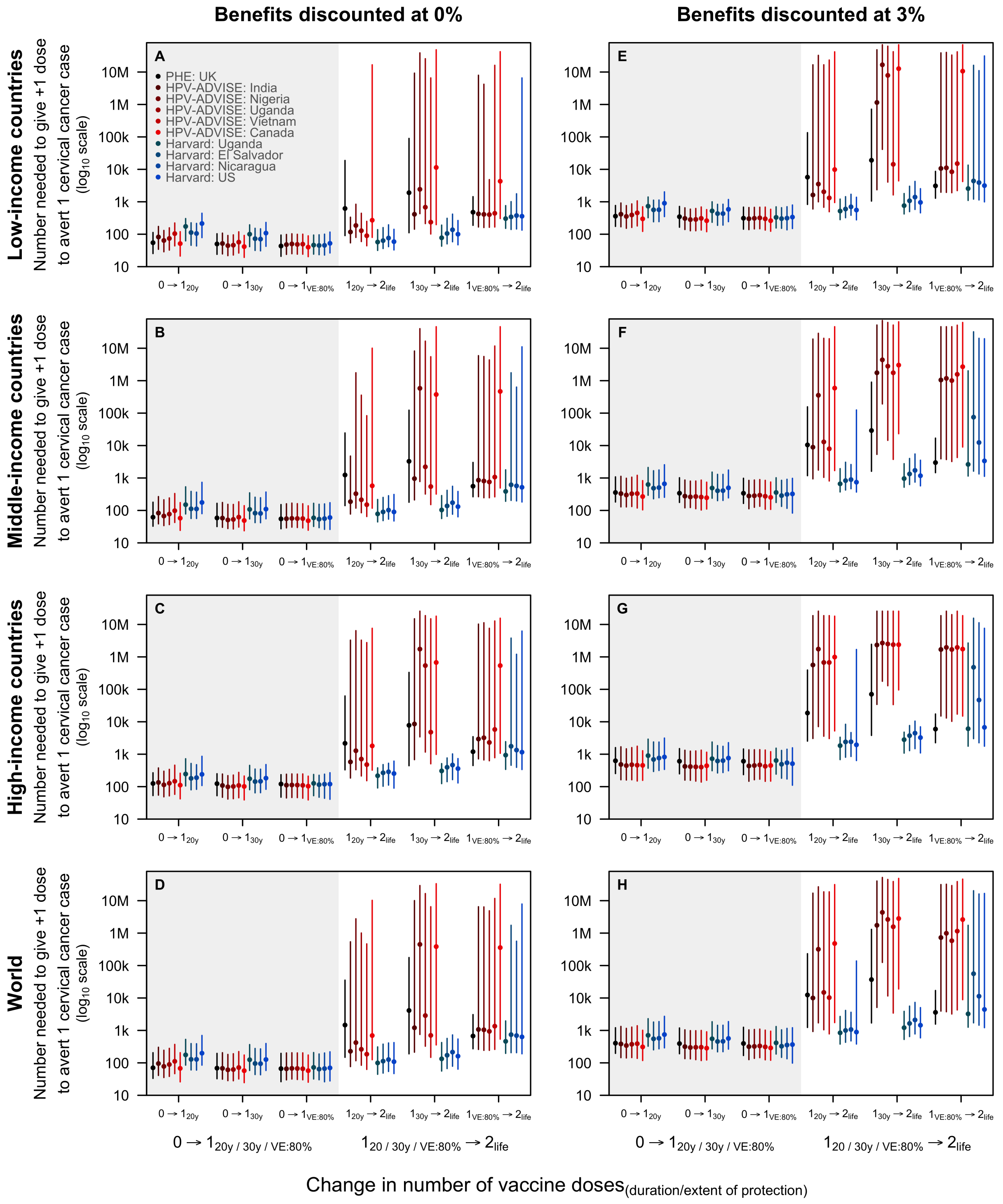

Figure 11: Number of girls needed to be vaccinated to avert one additional case over the years 2021–2120.
